# Population-based analysis of *POT1* variants in a cutaneous melanoma case-control cohort

**DOI:** 10.1101/2022.05.16.22274971

**Authors:** Irving Simonin-Wilmer, Raul Ossio, Emmett Leddin, Mark Harland, Karen A. Pooley, Mauricio Gerardo Martil de la Garza, Sofia Obolenski, James Hewinson, Chi C. Wong, Vivek Iyer, John C. Taylor, Julia A. Newton-Bishop, D. Timothy Bishop, G. Andrés Cisneros, Mark M. Iles, David J. Adams, Carla Daniela Robles-Espinoza

## Abstract

Pathogenic germline variants in the protection of telomeres 1 gene (*POT1*) have been associated with predisposition to a range of tumor types, including melanoma, glioma, leukemia and cardioangiosarcoma. We sequenced all coding exons of the *POT1* gene in 2,929 European-descent melanoma cases and 3,298 controls, identifying 43 protein-changing genetic variants. We performed POT1-telomere binding assays for all missense and stop gained variants, finding nine variants that impair or disrupt protein-telomere complex formation, and we further define the role of variants in the regulation of telomere length and complex formation through molecular dynamics simulations. We determine that *POT1* variants are a minor contributor to melanoma burden in the general population, with only about 0.5% of melanoma cases carrying germline pathogenic variants in this gene, but should be screened in individuals with a strong family history of melanoma and/or multiple malignancies.

## Introduction

Since the discovery of pathogenic alleles of *CDKN2A* twenty five years ago^1^, a number of other variants that increase melanoma risk have been uncovered by genome-wide association studies (GWAS)^2^ and the genomic analysis of melanoma-predisposed families. These variants affect biological pathways related to pigmentation (such as alleles of *MC1R, the “red hair” gene)*, naevus count, including genetic variation adjacent to *PLA2G6*, cell cycle and senescence, comprising changes in *CDKN2A* and *CDK4*, and telomere regulation^3^. Of note, pathogenic variants in the protection of telomeres gene (*POT1*) have been associated with melanoma, as well as other types of cancer such as glioma^4^, leukaemia^5^ and lymphoma^6^. As such, pathogenic germline *POT1* variants have recently been recognised as defining a novel tumor predisposition syndrome^7^. Genetic variation around *POT1* has also been found to be associated with melanoma in recent large-scale GWAS studies^8^.

*POT1* encodes a single-stranded DNA (ssDNA) binding protein that forms part of the shelterin complex, a group of proteins that have functions in telomere protection by allowing cells to distinguish the ends of chromosomes from sites of DNA damage, and also function to regulate telomere length^9^. In recent years, sequencing of melanoma-predisposed individuals has revealed a number of pathogenic alleles of *POT1* which affect the ability of POT1 to bind to ssDNA and therefore lead to longer and abnormal telomeres^10–12^. This, in turn, may promote carcinogenesis through the accumulation of damage at chromosome ends and a delay in the onset of cell senescence. Further, a recent study has identified *POT1* variants that lead to shorter telomeres^13^, emphasizing the need to identify and catalogue the consequences of these genetic changes in carriers.

As estimates have suggested that *POT1* may be the second major high-penetrance melanoma susceptibility gene after *CDKN2A*, being causal of disease predisposition in 2-4% of *CDKN2A/CDK4*-negative families^10, 14^, it has been included in multiple panels for genetic testing of melanoma families. As such, and in order to inform genetic counselling, there is a need to identify which genetic variants abrogate POT1 function leading to telomere dysregulation, as well as to determine their frequency in population-ascertained melanoma cases. In this study, we performed experimental and bioinformatic analyses to identify germline variants that disrupt the POT1-ssDNA complex and lead to telomere length alterations.

## Results

This study included 2,929 melanoma cases and 3,298 controls, making up a total of 6,227 European-descent (British) individuals from two distinct melanoma cohorts plus a population cohort (**Supplementary Methods**). We sequenced all *POT1* coding exons on the MiSeq platform. After alignment, variant calling and quality filtering, we identified 43 protein-altering variants in *POT1,* by Fluidigm PCR-based amplicon sequencing and validated them by target capture with Agilent SureSelect probes and Illumina sequencing (**Supplementary Methods**, **Figure 1**, **Supplementary Table 1**).

**Figure 1.**
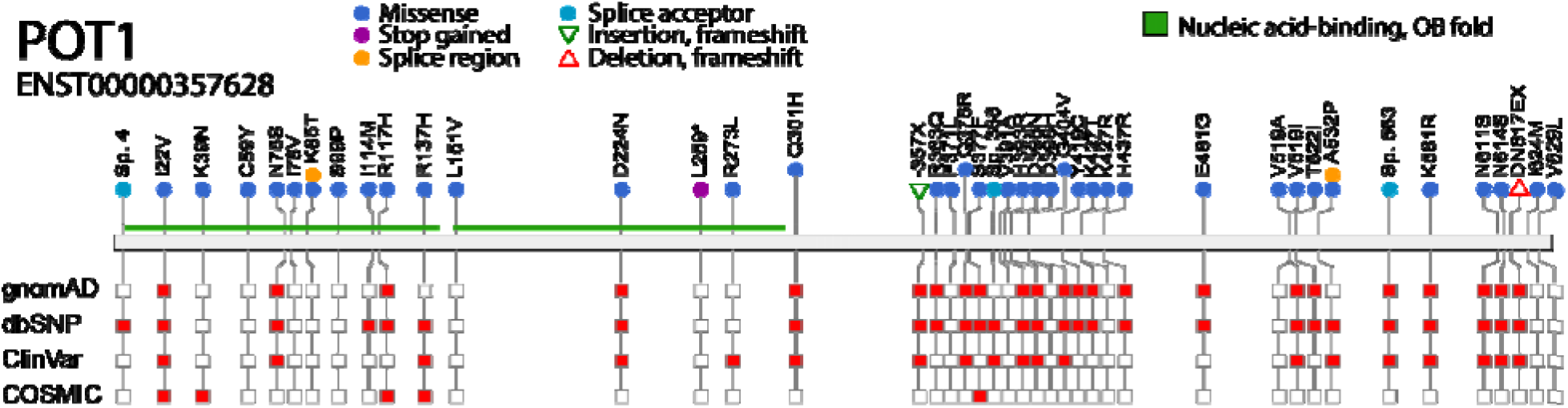
Schematic diagram of POT1 variants identified in this case-control cohort study. Variants are shown on the primary protein structure with their consequence (in a colored circle or triangle) and their presence (red square) or absence (empty square) in publicly available datasets (gnomAD v2.1, dbSNP build 151, ClinVar v86 and COSMIC v86). The OB domains are shown in green. For details on numbers of cases and controls see **Supplementary Table 1**. Figure created with VCF/Plotein [12].

In order to assess whether the detected variants impair telomere regulation, we analysed the ability of *in vitro*-translated POT1 proteins containing all missense and stop-gained variants (38/43 variants in total, **Supplementary Table 1**) to bind to a telomere-like oligo via EMSA experiments (**Supplementary Methods**). Our results indicate that four variants completely disrupted POT1-ssDNA complex formation (C59Y, R137H, L259X and R273L), whereas a further five appear to reduce the affinity of the interaction (K39N, K85T, S99P, R117H and D224N) (**Figure 2, Supplementary Figure 1**). Of note, as expected, all variants that altered POT1-DNA binding fall within the OB domains.

**Figure 2.**
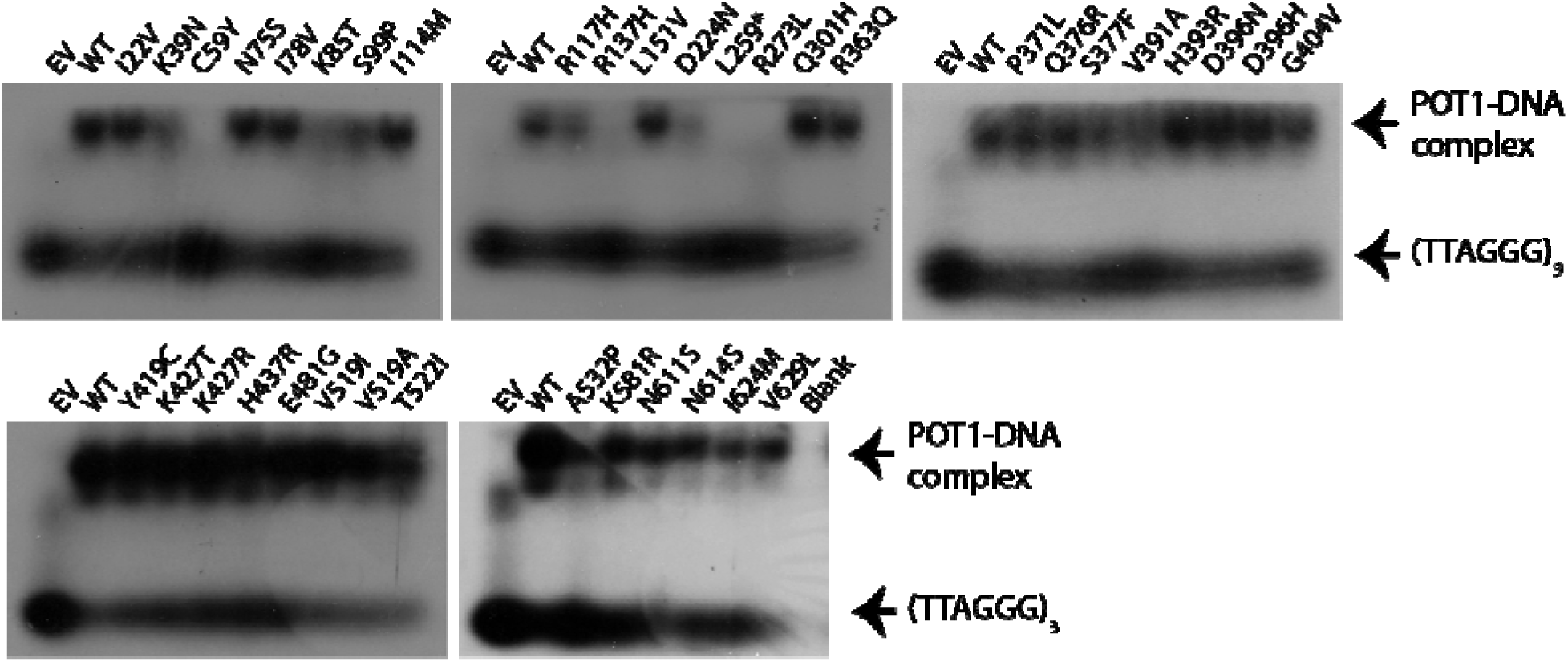
Electrophoretic mobility shift assays (EMSAs) testing the ability of in vitro- translated mutant POT1 proteins to bind a telomere-like oligo (TTAGGGTTAGGGTTAGGG). EV: Empty vector. WT: Wild type protein.

Variants were classified in three groups according to their pathogenicity: Group 1 variants were confirmed by EMSA to disrupt the POT1-DNA complex or were those strongly suspected as pathogenic (frameshift and splice acceptor variants). We included variants with reduced binding in this category due to their high conservation across species (**Supplementary Figure 2**) and prior evidence that these may be pathogenic (R117H^16^ and D224N^11^). In total, 14/43 variants were classified in this group, with 10 of these falling in the OB domains (**Supplementary Table 1**). Group 2 variants were those predicted deleterious and probably damaging by both the SIFT and PolyPhen algorithms, but that did not disrupt POT1-DNA binding (4/43 variants). These variants may impair the function of the protein in other ways. The remaining variants (25) were classified in Group 3. For group 1, 15 cases (0.51%) carried a variant, while 8 (0.24%) controls did (Two-tailed Fisher’s exact test, *P*-value = 0.095, odds ratio (OR) = 2.12). For Groups 1+2, 22 cases (0.75%) carried a variant, while 14 controls (0.42%) did (Two-tailed Fisher’s exact test, *P*-value 0.096, OR=1.78). Finally, for Group 3, 127 cases (4.3%) carried a variant as did 151 controls (4.6%) (Two-tailed Fisher’s exact test, *P*-value 0.66). Overall, while about twice as many cases as controls carried predicted pathogenic variants in *POT1*, this difference was not conventionally statistically significant likely because of limited power even with a study this size.

We next sought to determine whether the variants we detected had any effect on telomere regulation. For this, we measured telomere length in *POT1* variant carriers and non-carriers from the same populations (**Supplementary Methods**). After standardizing lengths by plate and adjusting them for cohort via a linear model (**Supplementary Table 2, Supplementary Figure 3**), we observed that only the individuals carrying the K39N (percentile 98 when compared to controls) and the R273L (percentile 99 when compared to controls) variants had telomeres that were substantially longer than the mean (**Figure 3**). We also observed that some individuals with splice variants or variants that showed reduced DNA binding also had telomeres on the longer side of the distribution (*e.g.* K85T, percentile 91, L259X, percentile 90, one individual carrying Sp. 388, percentile 97) but others did not (e.g. S99P, percentile 31, most individuals with variants in splice sites). Individuals with the D224N variant had telomere lengths scattered throughout the whole distribution in contrast to previous reports suggesting that these variants increase telomere length^11^ (**Figure 3**). It is worth noting that out of the 10 Leeds melanoma cases with Group 1 variants, five had early onset melanoma (<50 years of age) or a family history of the disease.

**Figure 3.**
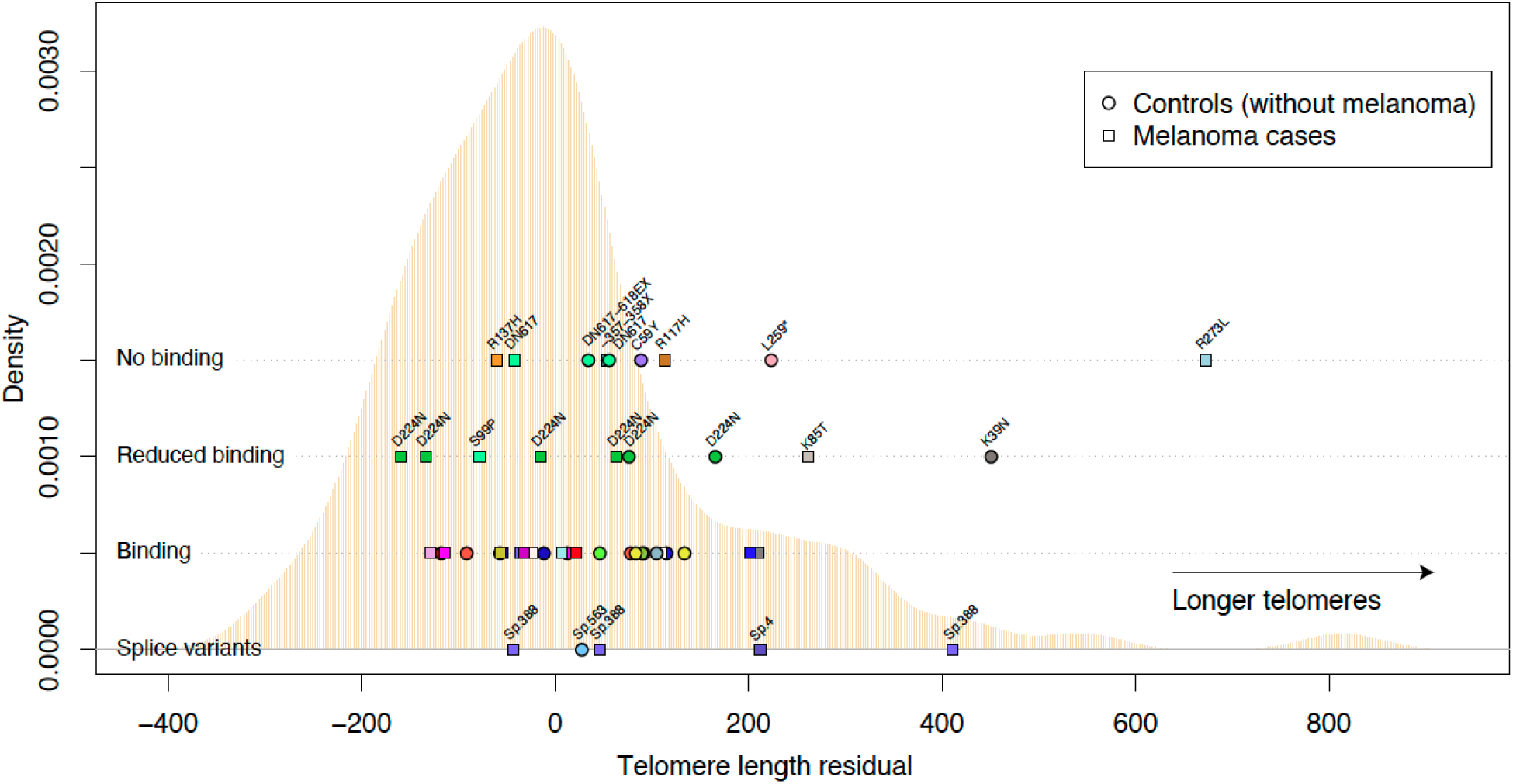
Telomere length of carriers of pathogenic *POT1* variants depicted over a telomere length distribution of melanoma cases and controls with no pathogenic *POT1* variants. The distribution of the means of residuals from the linear model distribution of telomere lengths for individuals with no *POT1* variants is depicted in beige. The mean of the adjusted telomere lengths for individuals with *POT1* variants are shown on top according to the variant type (No binding, reduced binding or binding according to EMSA, and splice variants). Melanoma cases are shown in squares and controls in circles. Each variant is shown in a different color. For the ‘Binding’ row, the variants from left to right are: P371L, I624M, N611S, K427T, D396H, V629L, H393R, L151V, N75S, H393R, K581R, E481G, S377F, N614S, Y419C- G404V, D396N, I78V, V519A, T522I, I114M, R363Q-V391A, K427R, H437R, V519I, H393R, A532P.

Because the K39N, C59Y, and D224N variants are found in controls and show POT1-ssDNA complex disruption, we further investigated them using molecular dynamics simulations (**Supplementary Methods**). Our results suggest that all three mutations affect the dynamics of the system when compared to the WT structure, as evidenced by the first and second normal modes (**Supplementary Figure 4A–H**, **Supplementary Movie**). Existing protein structures for *POT1* also imply that there are conformational differences between the *POT1*-ssDNA and *POT1*-ACD structures^17–20^. As a result, the structural differences noted within the *POT1* mutants investigated here may affect shelterin complex formation, but further investigation is necessary. Additional analyses of root mean square deviation (RMSD), root mean square fluctuation (RMSF), residue-wise correlations, secondary structure, energy decomposition analysis, and hydrogen bond interactions are all consistent with the computational results reported herein (**Supplementary Figures 4I-L, 5-17, Supplementary Tables 3-7**).

## Discussion

Even though *POT1* seems to be the second major melanoma susceptibility gene, with 2-4% of *CDKN2A/CDK4*-wild type families carrying a pathogenic variant in this gene, its contribution to melanoma risk burden in the general population is minor, with only about 0.5% of cases carrying pathogenic variants. Telomere length calculations confirm known associations of variants with longer telomeres (R273L^10^, R117H^11, 16^) and found associations with other pathogenic variants (K39N, K85T, and confirmation of longer telomere length for A532P, percentile 93, a variant originally reported in Ref. ^11^), but for other variants the association with length was not clear (for example, all three carriers of Sp. 388 and six of D224N had telomere lengths scattered throughout the distribution). Although a prior study had shown slightly longer telomeres for carriers of R117H^11^, the carrier melanoma case in this cohort had normal-length telomeres. This may reflect the many mechanisms, including other genetic variants and lifestyle, by which telomere length can be affected. Telomere length for some control individuals (without reported melanoma) with pathogenic variants (*e.g.*, K39N and both controls carrying D224N) also showed an increase in telomere length, which may portend an increased risk of tumorigenesis in these individuals or indicate that other factors are necessary for melanoma genesis.

Although in this study we have attempted to identify pathogenic *POT1* variants through DNA-binding assays, the function of proteins with variants outside the OB domains may also be compromised by other mechanisms. For example, another study has concluded that the *POT1* A532P variant shows impaired ACD binding, which may also lead to telomere dysregulation^22^. Therefore, further systematic experiments are needed to address other *POT1* functions to provide a more complete catalogue of variants that alter protein function and therefore that lead to cancer predisposition.

## Materials and methods

### Participants and sample collection

Samples used in this study came from three different cohorts. The Leeds Melanoma Case Control Study has recruited population-ascertained melanoma cases and the same sex and 5-year age group controls predominantly from the Yorkshire, UK geographical area since the year 2000 (NRES Committee North East - Northern and Yorkshire, MREC/01/3/057) [16]. Additionally, samples were included from the Study of Epidemiology and Risk Factors in Cancer Heredity (SEARCH) series of population-based studies in Eastern England (Cambridgeshire South Research Ethics Committee, 05/MRE05/1) [17]. Finally, controls were supplemented with samples from the Wellcome Trust Case Control Consortium [18] (South East Multicentre Research Ethics Committee, 05/Q0106/74) (**Supplementary Table 8**). DNA from whole blood was extracted for exon capture and sequencing.

### Sequencing and variant calling

We used Fluidigm PCR-based amplicon sequencing to amplify all of the coding exons and splice junctions of the *POT1* gene in 7,024 samples and sequenced these products on the MiSeq platform. After aligning with BWA and filtering to keep only highly covered samples (Those with >94% of coding *POT1* exon bases covered >= 10 reads with MQ >= 50 and base quality >= 20), keeping only one sample out of each pair of relatives, removing samples with non-European ancestry and samples from patients that withdrew from the study, and removing related controls, 6,227 samples remained. These samples included 2,929 cases (1,574 from the Leeds Case Control Consortium and 1,355 from the SEARCH Consortium) and 3,298 controls (1,431 from the WTCCC collection, 459 from the Leeds Case Control Consortium and 1,408 from the SEARCH Consortium) (**Supplementary Table 8**). We took forward for analysis the union of calls made by HaplotypeCaller (command line below in this section) and Samtools mpileup (parameters -t DP, SP -C50 -m2 -F0.0005 -d 10000 -ug), and performed quality variant filtering (mpileup: QUAL>=20 && (DP4[2]>30 || DP4[3]>30); GATK: QUAL>=20 && AD[0:1]>30). We also removed indel calls between GRCh37 coordinates 7:124475296-124475328, as this highly repetitive region (at an intron-exon boundary) seems to be germline microsatellite unstable and may be prone to false positive calls. We called 141 different protein-altering variants (missense, stops, frameshifts and splice acceptor/donor sites) in 3,546 samples electing to use a low stringency approach so as to capture all possible variants for validation. Validation by re-sequencing with Sanger or Illumina technologies was performed for at least one sample for all detected variants, often re- sequencing all carrier samples (**Supplementary Table 9**). Illumina sequencing was performed by exome capture with Agilent Technologies probes, using the WTSI v4 Solid Tumor Panel which included other established melanoma predisposition genes such as *CDKN2A*, *BAP1* and *CDK4*. Sequencing captured all exons and exon/intron boundaries of *POT1*, and succeeded for 164 samples. Overall, 158/164 samples were covered to an average depth higher than 10x across all *POT1* coding exons (**Supplementary Table 10**). Variants were called from these data using Samtools mpileup in pooled mode. By applying this approach, we validated 42 unique variants originally identified using the Fluidigm PCR-based amplicon but importantly identified no new variant positions by this method. Capillary resequencing of variants found in the 19 samples that failed library preparation or were included in a later sequencing effort confirmed one additional variant, for a total of 43/141 protein-altering variants confirmed (**Supplementary Table 1, Supplementary Table 11**). No additional variants were called in the resequenced samples. Consequences were predicted with the Variant Effect Predictor (VEP), from Ensembl release 104, using the web tool with the GRCh37 human reference genome. Since our exon capture sequenced additional known melanoma driver genes (above) we screened all samples found to carry mutations in *POT1* to exclude the possibility that they also carried pathogenic variants in drivers *CDK4*, *CDKN2A* and *BAP1*. Three protein-coding variants were found in *CDKN2A* in *POT1* variant carriers, although these *POT1* variants were classified as benign according to our G-strand binding assays (**Supplementary Table 12**, reference transcripts *CDKN2A*: ENST00000304494, *CDK4*: ENST00000257904, *BAP1*: ENST00000460680).

### *In vitro* translation and G strand binding assays

pEX-POT1 plasmid vectors, harboring wild-type or mutant POT1 ORF sequences downstream of a T7 promoter, were used for in vitro translation reactions with TNT coupled reticulocyte lysate kit (Promega) according to the manufacturer’s instructions. Protein expression was verified by immunoblotting an aliquot of each reaction with anti-POT1 antibodies (Abcam, ab124784). A telomeric oligonucleotide probe (GGTTAGGGTTAGGGTTAGGG) was end-labelled using [-32P] ATP (Perkin Elmer) with T4 polynucleotide kinase (New England Biolabs). Unincorporated nucleotides were removed using illustra MicroSpin G-25 columns (GE Healthcare) according to the manufacturer’s instructions. DNA-binding assays were performed by mixing 5µl translation reaction in 20 µl final volume containing binding buffer (25 mM HEPES-NaOH [pH 7.5], 100 mM NaCl, 1 mM EDTA and 5% glycerol), 1 µg poly (dI-dC) (Thermo) and 10 nM [32P]-labelled telomeric oligonucleotide probe for 10 min at room temperature. Reactions were separated on 6% DNA retardation gels (Novex) in 0.5 TBE buffer at 80 V. Gels were dried and exposed to Hyperfilm MP film (Amersham), which was developed using a Compact X4 machine (Xograph).

### Analysis of telomere length by telomere repeat PCR

We measured telomere length in all Illumina re-sequenced cases and controls from the Leeds, SEARCH and WTCC cohorts who carried a potential *POT1* variant according to the initial Fluidigm analysis, as well as age and sex-matched controls (A total of 174 samples (of which 66 belong to the Leeds cohort, 86 to SEARCH and 22 to WTCC; 105 are melanoma cases and 69 are non-melanoma controls, and 49 pathogenic POT1 variant carriers and 130 non-carriers)). Telomere length was quantified by real-time PCR using the ‘Absolute Human Telomere Length Quantification qPCR Assay Kit (AHTLQ)’ (ScienCell Research Laboratories, CA, USA) according to the manufacturer’s instructions. Each DNA sample was amplified in two separate reactions: using the telomere (TEL) primer set; and the single copy reference (SCR) primer set. The telomere primer set recognises and amplifies telomere sequences. The SCR primer set amplifies a 100bp region on chromosome 17, and acts as a reference for normalisation.

Reactions were carried out in 20 μL volume: 1 μL DNA (5ng); 2 μL primer (TEL or SCR); 10 μL 2×GoldNStart TaqGreen qPCR master mix; and 7 μL nuclease-free water. A QuantStudio 5 Real Time PCR machine (Life Technologies, CA, USA) was used for qPCR, using a 96-well plate format. The PCR conditions were: initial denaturation at 95°C for 10 minutes, then 32 cycles of 95°C for 20 seconds, 52°C for 20 seconds and 72°C for 45 seconds. All reactions were performed in triplicate, and the same reference genomic DNA sample was included in each run. Data were analysed using the QuantStudio Design and Analysis Software version 1.4.1. (Life Technologies, CA, USA), and absolute telomere length was calculated by reference to the DNA standard using comparative ddCq according to the AHTLQ kit instructions.

A linear model adjusting for age at diagnosis, sex and cohort was done with the individuals that did not have pathogenic variants (pathogenic variants were defined as all detected variants except for I22V, Q301H, Q376R, and G404V, which all have a gnomAD overall allele frequency higher than 1 x 10^-4^), whether melanoma cases or controls. The linear model showed that neither age nor sex where significantly related to the telomere length in our data. There is probably too much noise introduced by cohort origin, so we opted for using only cohort to control our data. The residuals of this linear model were used to create a telomere length distribution for this cohort. Telomere length adjustment for pathogenic variant carrier individuals was done separately with the same parameters calculated from the population distribution.

### Molecular dynamics simulations of WT and variant POT1-ssDNA

Molecular dynamics (MD) simulations were performed using AMBER20’s pmemd.cuda with the ff19SB force field for protein and OL15 for DNA^23–26^. The model was constructed from PDB ID 1XJV,^19, 20^ using *Coot* and MolProbity to alleviate bad clashes^27, 28^. We used the Modeller interface in UCSF Chimera to incorporate missing residues,^29–31^ as well as Chimera’s integrated Dunbrack rotamer library to create the K39N, C59Y, and D224N variants^32^. The systems were solvated with TIP3P water in a cube that extended 12 Å from the complex surface, and potassium was added to neutralize the charge of the system^33^. All MD simulations were run in triplicate for 250 ns while holding the number of atoms, pressure, and temperature constant (NPT ensemble) with the Langevin thermostat and barostat. A 9 Å cutoff was used for long-range non-bonded interactions with the smooth particle mesh Ewald method for electrostatics^34^. AMBER’s cpptraj program was used for root mean square deviation (RMSD), root mean square fluctuation (RMSF), hydrogen bond interactions, and secondary structure analyses^35^. Both cpptraj and the ProDy module in VMD were used for normal mode analysis^36, 37^. A Fortran90 program developed by the Cisneros group was used to perform an energy decomposition analysis (EDA) on each simulation^38^. The data.table, tidyverse, and abind packages of R were used to analyze the EDA data and hydrogen bond interactions^39–42^. VMD, UCSF Chimera, gnuplot, matplotlib, and ggplot2 were used for data visualization and image creation^31, 37, 43–45^.

### Command lines used

GATK HaplotypeCaller command line:

analysis_type=HaplotypeCaller input_file=[example.bam] showFullBamList=false read_buffer_size=null phone_home=NO_ET gatk_key=gatk.key tag=NA read_filter=[] disable_read_filter=[] intervals=[pot1.bed, 7:1-159138663] excludeIntervals=null interval_set_rule=INTERSECTION interval_merging=ALL interval_padding=0 reference_sequence=hs37d5.fa nonDeterministicRandomSeed=false disableDithering=false maxRunti me=-1 maxRuntimeUnits=MINUTES downsampling_type=BY_SAMPLE downsample_to_fraction=null downsample_to_coverage=500 baq=OFF baqGapOpenPenalty=40.0 refactor_NDN_cigar_string=false fix_misencoded_quality_scores=false allow_potentially_misencoded_quality_scores=false useOriginalQualities=false defaultBaseQualities=-1 performanceLog=null BQSR=null quantize_quals=0 disable_indel_quals=false emit_original_quals=false preserve_qscores_less_than=6 globalQScorePrior=-1.0 validation_strictness=SILENT remove_program_records=false keep_program_records=false sample_rename_mapping_file=null unsafe=null disable_auto_index_creation_and_locking_when_reading_rods=false no_cmdline_in_header=false sites_only=false never_trim_vcf_format_field=false bcf=false bam_compression=null simplifyBAM=false disable_bam_indexing=false generate_md5=false num_threads=1 num_cpu_threads_per_data_thread=1 num_io_threads=0 monitorThreadEfficiency=false num_bam_file_handles=null read_group_black_list=null pedigree=[] pedigreeString=[] pedigreeValidationType=STRICT allow_intervals_with_unindexed_bam=false generateShadowBCF=false variant_index_type=DYNAMIC_SEEK variant_index_parameter=-1 logging_level=ERROR log_to_file=null help=false version=false out=/example_path/1_gatk_haplotype_caller_with_genome_chunking/7_1-159138663.gatk_haplotype.vcf.gz likelihoodCalculationEngine=PairHMM heterogeneousKmerSizeResolution=COMBO_MIN dbsnp=(RodBinding name= source=UNBOUND) dontTrimActiveRegions=false maxDiscARExtension=25 maxGGAARExtension=300 paddingAroundIndels=150 paddingAroundSNPs=20 comp=[] annotation=[ClippingRankSumTest, DepthPerSampleHC] excludeAnnotation=[] debug=false useFilteredReadsForAnnotations=false emitRefConfidence=NONE bamOutput=null bamWriterType=CALLED_HAPLOTYPES disableOptimizations=false annotateNDA=false heterozygosity=0.001 indel_heterozygosity=1.25E-4 standard_min_confidence_threshold_for_calling=4.0 standard_min_confidence_threshold_for_emitting=4.0 max_alternate_alleles=6 input_prior=[] sample_ploidy=2 genotyping_mode=DISCOVERY alleles=(RodBinding name= source=UNBOUND) contamination_fraction_to_filter=0.0 contamination_fraction_per_sample_file=null p_nonref_model=null exactcallslog=null output_mode=EMIT_VARIANTS_ONLY allSitePLs=false gcpHMM=10 pair_hmm_implementation=VECTOR_LOGLESS_CACHING pair_hmm_sub_implementation=ENABLE_ALL always_load_vector_logless_PairHMM_lib=false phredScaledGlobalReadMismappingRate=45 noFpga=false sample_name=null kmerSize=[10, 25] dontIncreaseKmerSizesForCycles=false allowNonUniqueKmersInRef=false numPruningSamples=1 recoverDanglingHeads=false doNotRecoverDanglingBranches=false minDanglingBranchLength=4 consensus=false maxNumHaplotypesInPopulation=128 errorCorrectKmers=false minPruning=2 debugGraphTransformations=false allowCyclesInKmerGraphToGeneratePaths=false graphOutput=null kmerLengthForReadErrorCorrection=25 minObservationsForKmerToBeSolid=20 GVCFGQBands=[1, 2, 3, 4, 5, 6, 7, 8, 9, 10, 11, 12, 13, 14, 15, 16, 17, 18, 19, 20, 21, 22, 23, 24, 25, 26, 27, 28, 29, 30, 31, 32, 33, 34, 35, 36, 37, 38, 39, 40, 41, 42, 43, 44, 45, 46, 47, 48, 49, 50, 51, 52, 53, 54, 55, 56, 57, 58, 59, 60, 70, 80, 90, 99] indelSizeToEliminateInRefModel=10 min_base_quality_score=10 includeUmappedReads=false useAllelesTrigger=false doNotRunPhysicalPhasing=true keepRG=null justDetermineActiveRegions=false dontGenotype=false dontUseSoftClippedBases=false captureAssemblyFailureBAM=false errorCorrectReads=false pcr_indel_model=CONSERVATIVE maxReadsInRegionPerSample=10000 minReadsPerAlignmentStart=10 mergeVariantsViaLD=false activityProfileOut=null activeRegionOut=null activeRegionIn=null activeRegionExtension=null forceActive=false activeRegionMaxSize=null bandPassSigma=null maxProbPropagationDistance=50 activeProbabilityThreshold=0.002 min_mapping_quality_score=20 filter_reads_with_N_cigar=false filter_mismatching_base_and_quals=false filter_bases_not_stored=false

VEP command:

./vep --af --af_gnomad --appris --biotype --buffer_size 500 --check_existing --distance 5000 --mane --polyphen b --pubmed -- regulatory --sift b --species homo_sapiens --symbol --transcript_version --tsl --cache --input_file [input_data] --output_file [output_file] --port 3337

## Supporting information

STROBE checklist

## Data Availability

Sequencing data for this study has been deposited in the European Genome-Phenome Archive, under study ID EGAS00001001964.

## Acknowledgments

We are deeply grateful to the patients and families that kindly donated the samples used in this study. We are thankful to Dr Charles Mein, Centre Manager of Barts and the London Genome Centre, for support during the initial phase of this project. The authors also wish to thank Jair S. García-Sotelo, Alejandro de León, Carlos S. Flores, and Luis A. Aguilar of the Laboratorio Nacional de Visualización Científica Avanzada from the National Autonomous University of Mexico, and Alejandra Castillo, Carina Díaz, Abigayl Hernández, and Eglee Lomelin of the International Laboratory for Human Genome Research, UNAM. I.S.-W. is a PhD student from Programa de Doctorado en Ciencias Biomédicas, Universidad Nacional Autónoma de México (UNAM). This work forms part of his dissertation.

## Authors’ contributions

I.S.-O.: sequencing and telomere length data analysis, R.O.: sequencing data analysis, E.L.: molecular dynamics simulation analysis, M.H.: qPCR assays and sample management, K.A.P.: qPCR assays and sample management, M.G.M.G.: molecular dynamics simulation analysis, S.O.: data analysis, J.H.: sample management, C.C.W.: telomere binding assays, V.I.: sequence variant calling, J.C.T.: sample management, J.A.N.-B.: patient management, manuscript writing, D.T.B.: statistical analysis supervision, manuscript writing, G.A.C.: molecular dynamics simulation analysis, manuscript writing, M.M.I.: statistical analysis supervision, D.J.A.: conceived and supervised study, manuscript writing, C.D.R.-E.: conceived and supervised study, sequencing and variant data analysis, manuscript writing.

## Supplementary Figures and legends

**Supplementary Figure 1.**
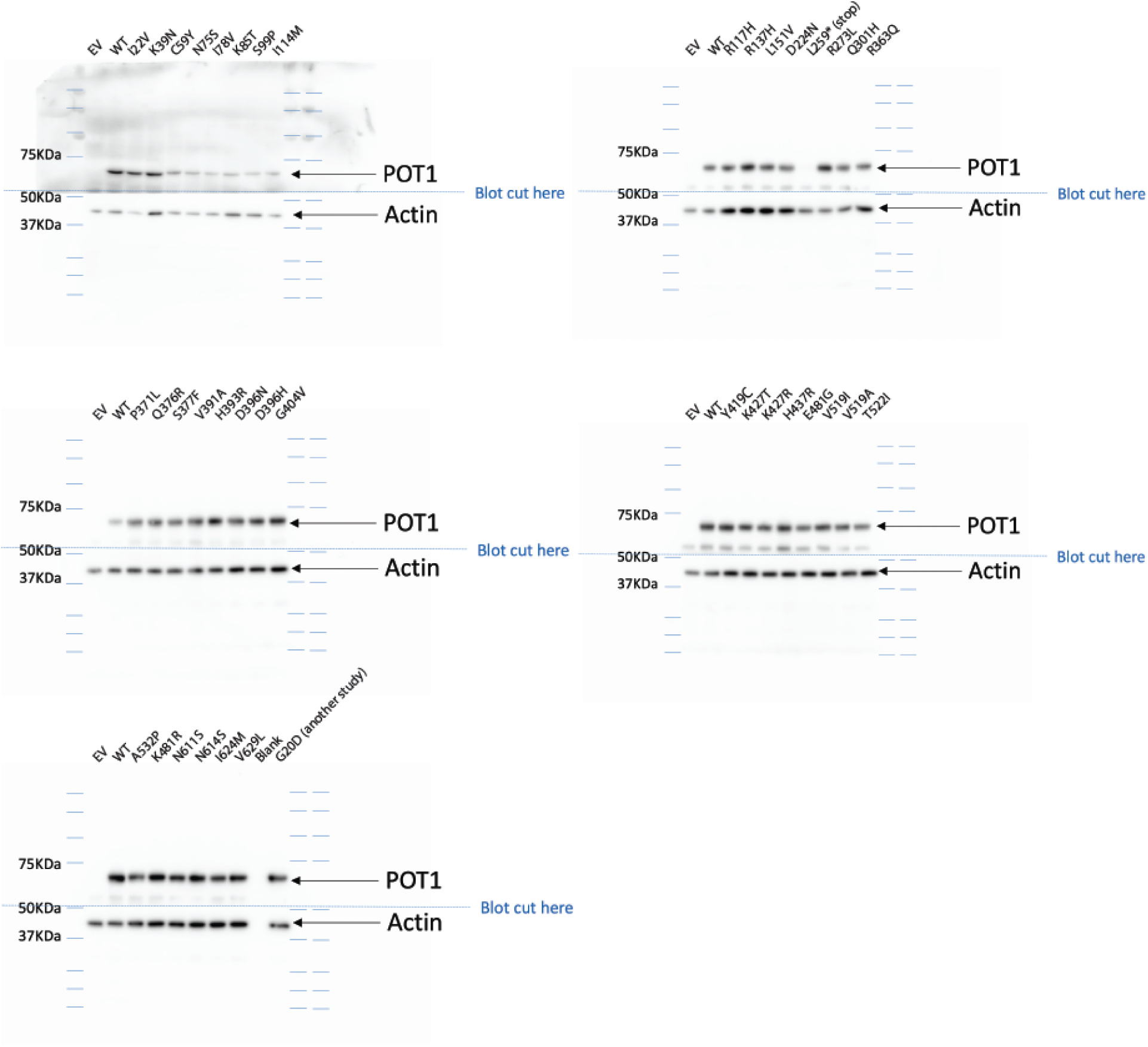
Expression control for the POT1-ssDNA binding assays. The POT1 protein is shown alongside the expression of actin in each reticulocyte lysate reaction. The gels are shown in the same order as those in Figure 2.

**Supplementary Figure 2.**
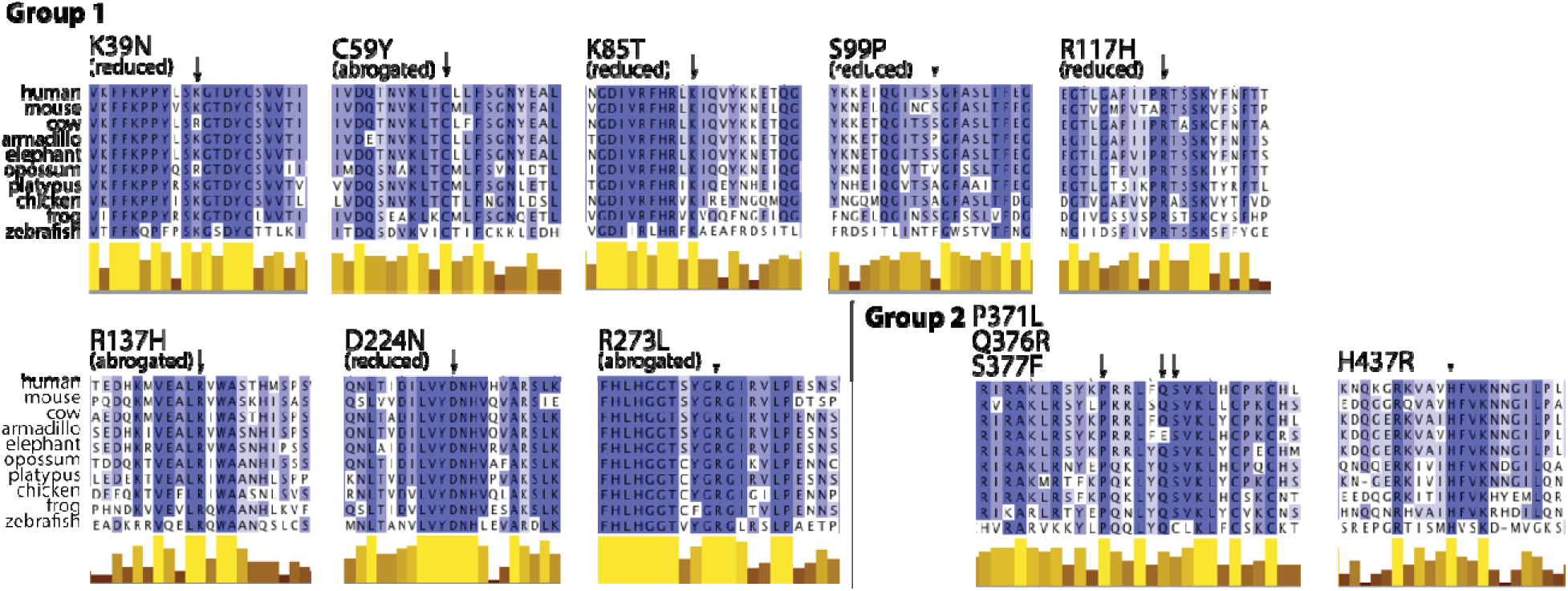
Multi-species alignment of the POT1 protein sequence fo missense variants in Pathogenicity Groups 1 and 2. A darker shade in an amino acid indicates higher sequence conservation across species. At the bottom, the height and color of the bars indicate sequence conservation level (taller and lighter bars indicate higher conserved residues). Protein sequences were downloaded from NCBI and are: NP_056265.2 (human), NP_598692.1 (mouse), DAA30462.1 (cow), XP_004478310.1 (armadillo), XP_003407293. (elephant), XP_007504312.1 (opossum), XP_001508179.2 (platypus), NP_996875.1 (chicken), AAI71328.1 (frog) and ADY16707.1 (zebrafish). Alignments were done with CLUSTAL O v 1.2.4 and rendered with Jalview.

**Supplementary Figure 3.**
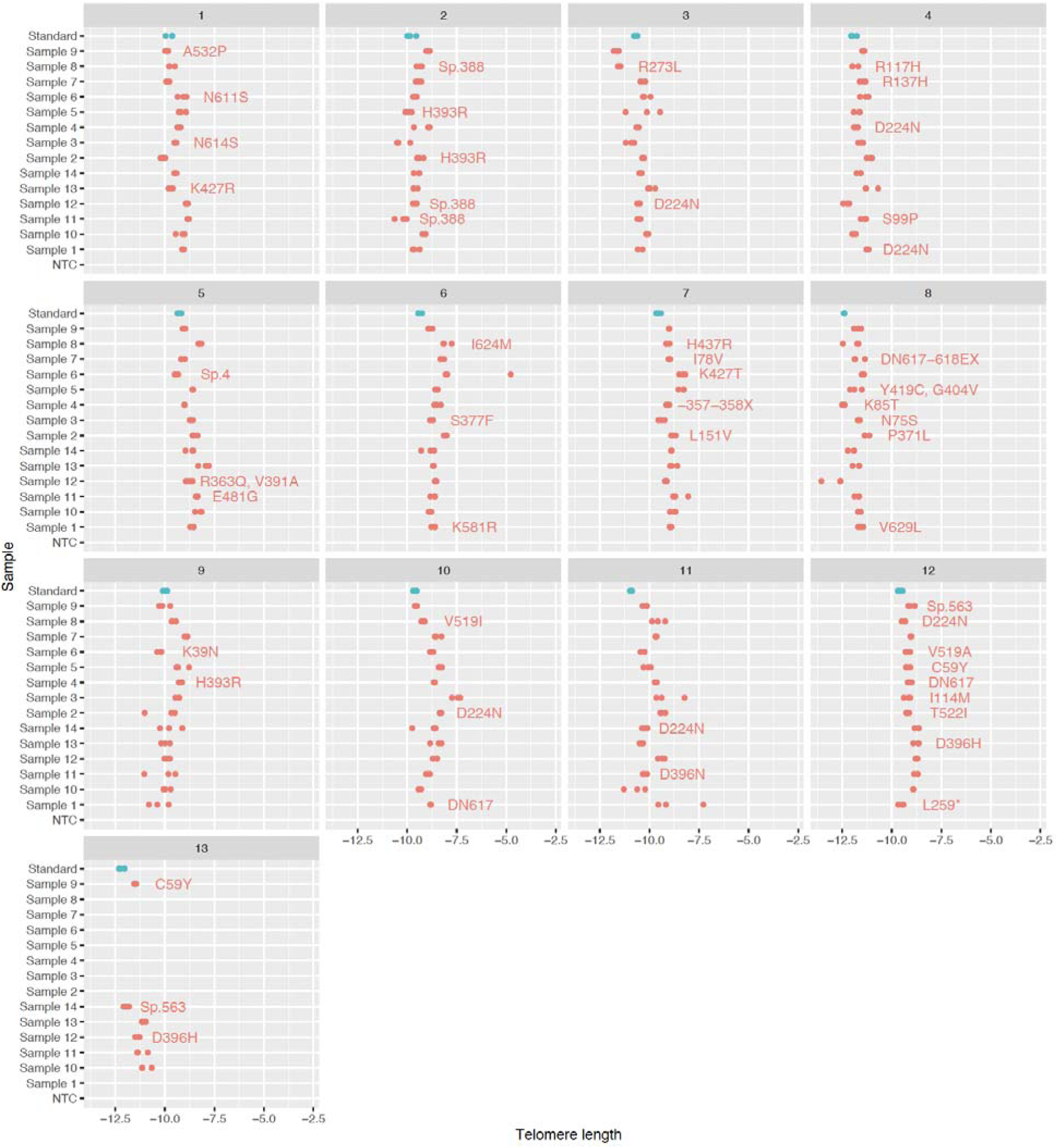
Raw telomere length measurements. Each plate is shown, with the standard of known telomere length shown in the top row. Three replicates per sample were measured and are shown, the mean of measurements was used for the linear model and the distribution seen in Figure 3. Variants carried by each sample are depicted.

**Supplementary Figure 4.**
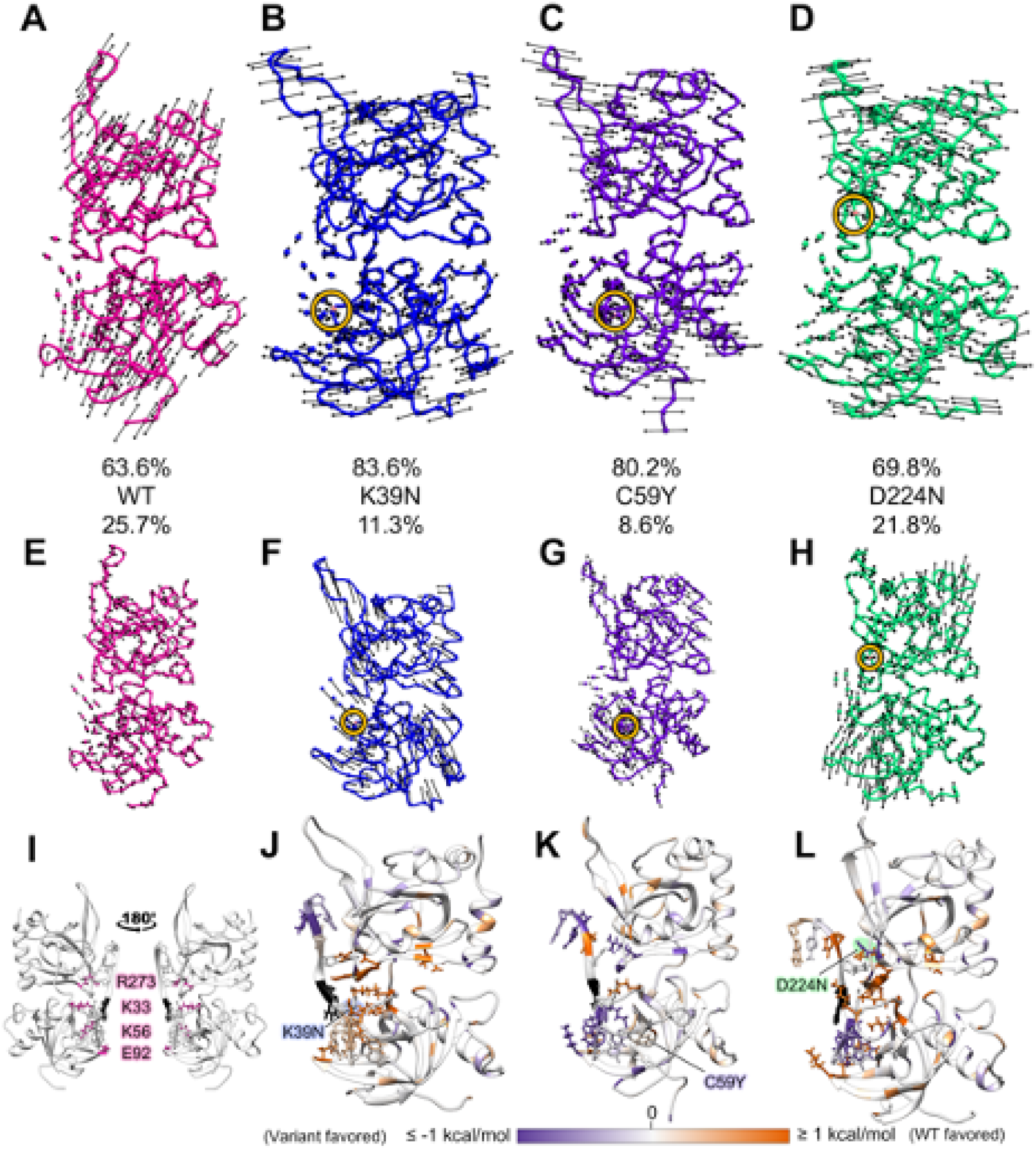
Variant differences in dynamics and non-bonded interactions. (A–D) First and (E–H) second normal modes for (A,E) WT (B, F) K39N (C, G) C59Y and (D, H) D224N. The mutation position is colored pink and circled in yellow. (I) EDA results show that K33, K56, E92, and R73 (pink) have different non-bonded interactions with dG6 (black) across all variants when compared to WT. (J–L) Differences in the total interaction energy (Coulomb and van der Waals) with respect to the dG6 position for (J) K39N – WT, (K) C59Y – WT, and (L) D224N – WT. Interactions favored in WT are colored orange, those favored in the variant are colored, purple, and the dG6 position is colored black. All data are mapped onto the respective variant structure.

**Supplementary Figure 5.**
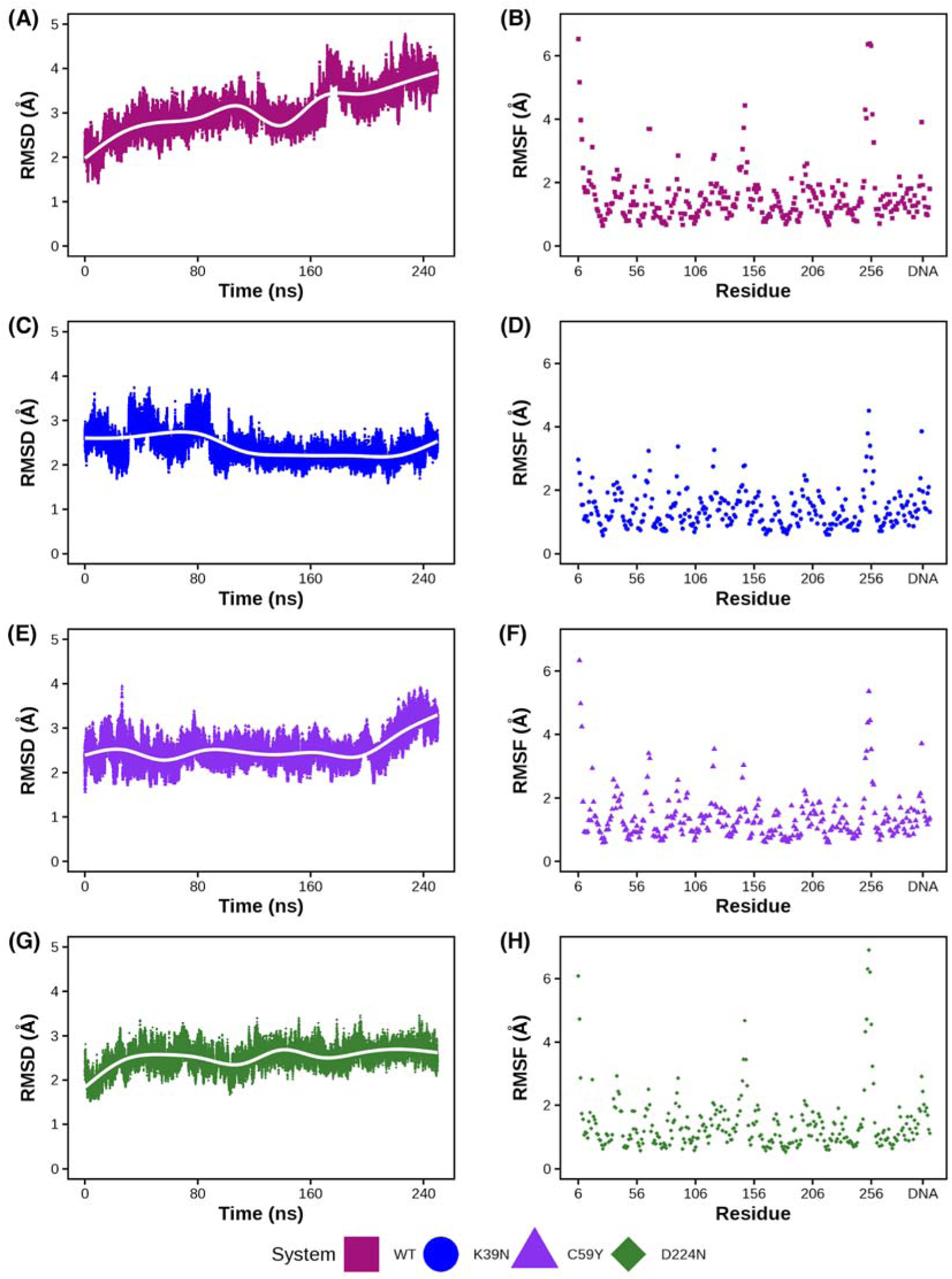
RMSD and RMSF. RMSD (left) and RMSF (right) of the first MD replicate.

**Supplementary Figure 6.**
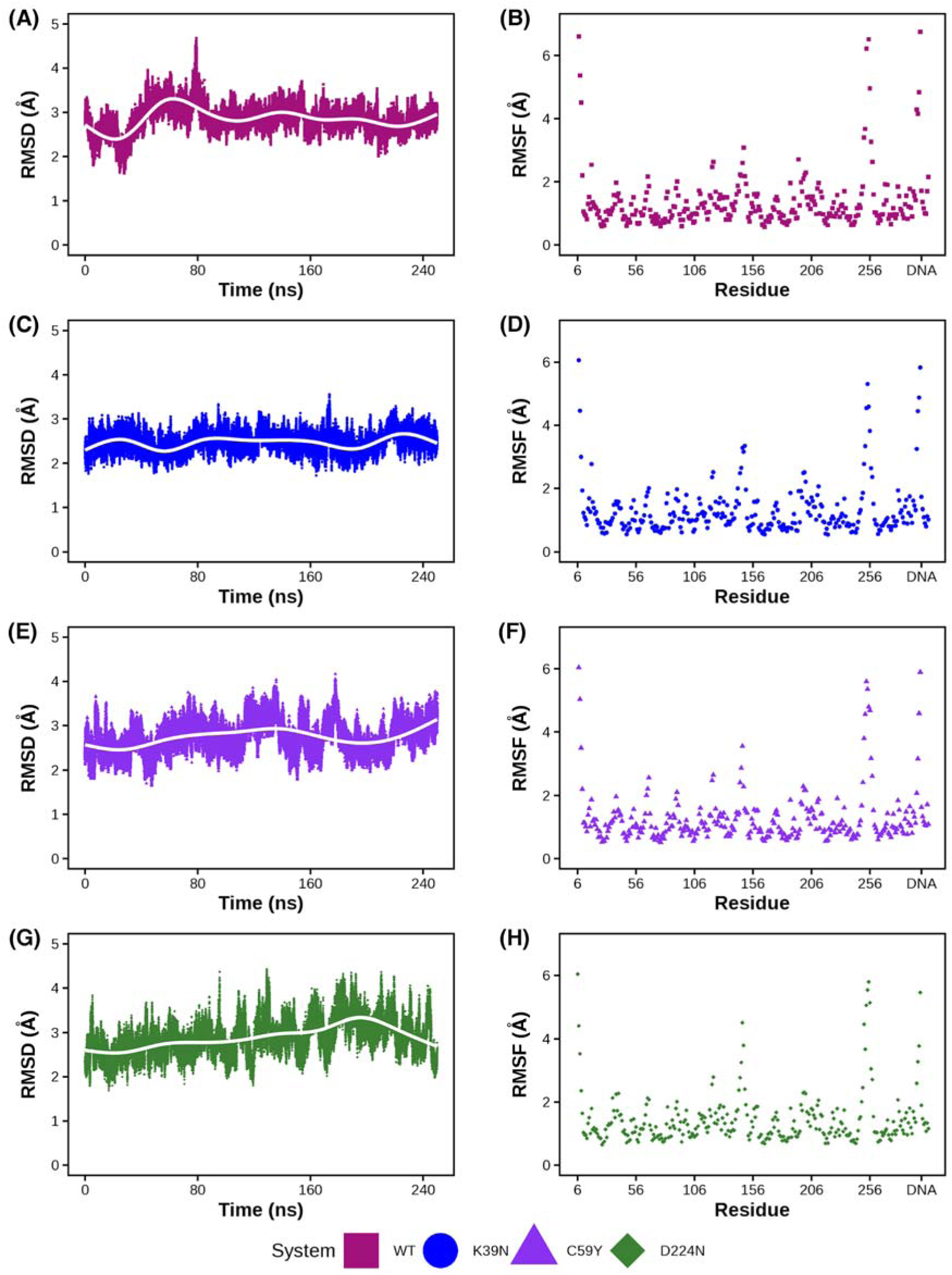
RMSD and RMSF. RMSD (left) and RMSF (right) of the second MD replicate.

**Supplementary Figure 7.**
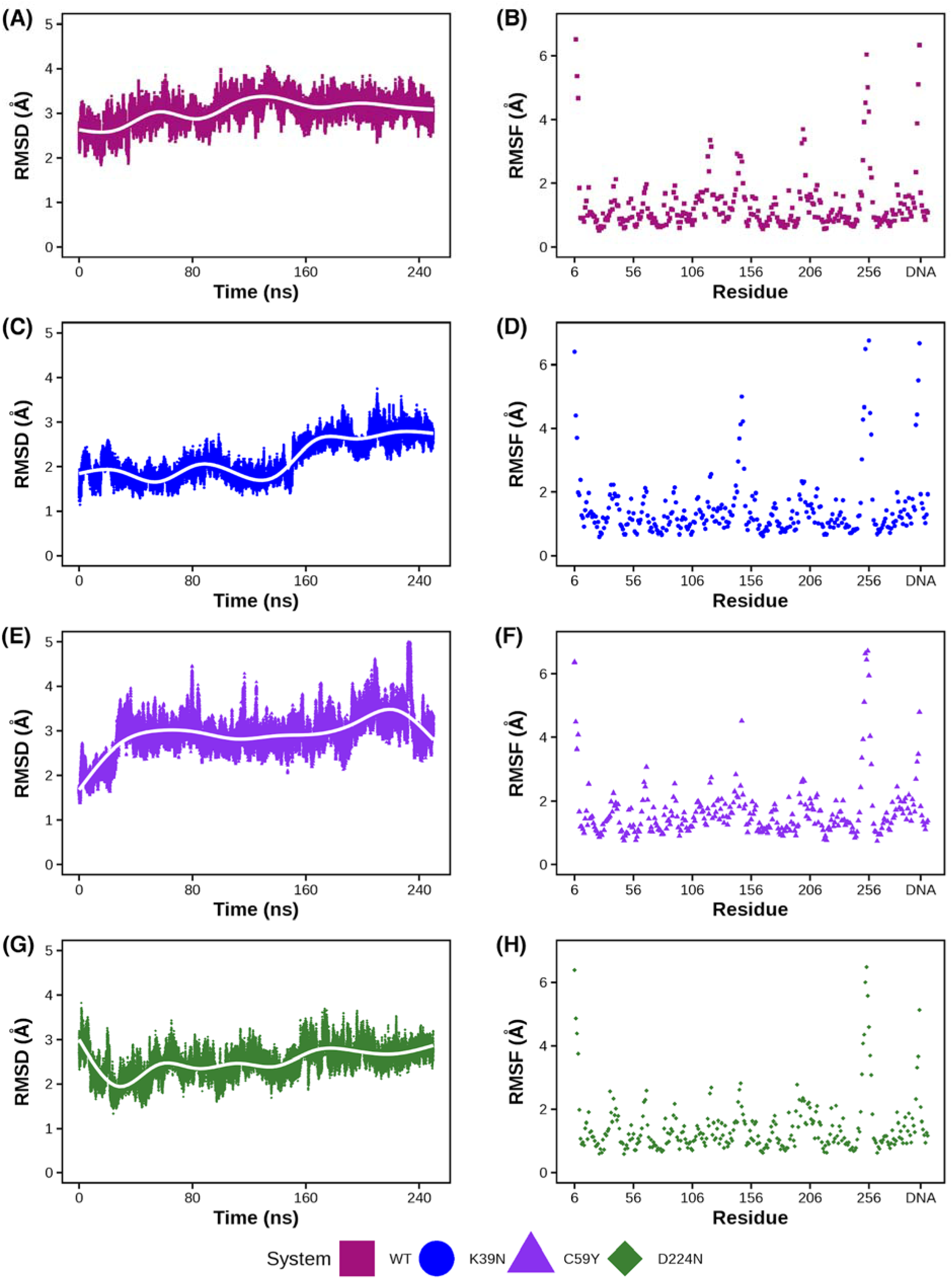
RMSD and RMSF. RMSD (left) and RMSF (right) of the third MD replicate.

**Supplementary Figure 8.**
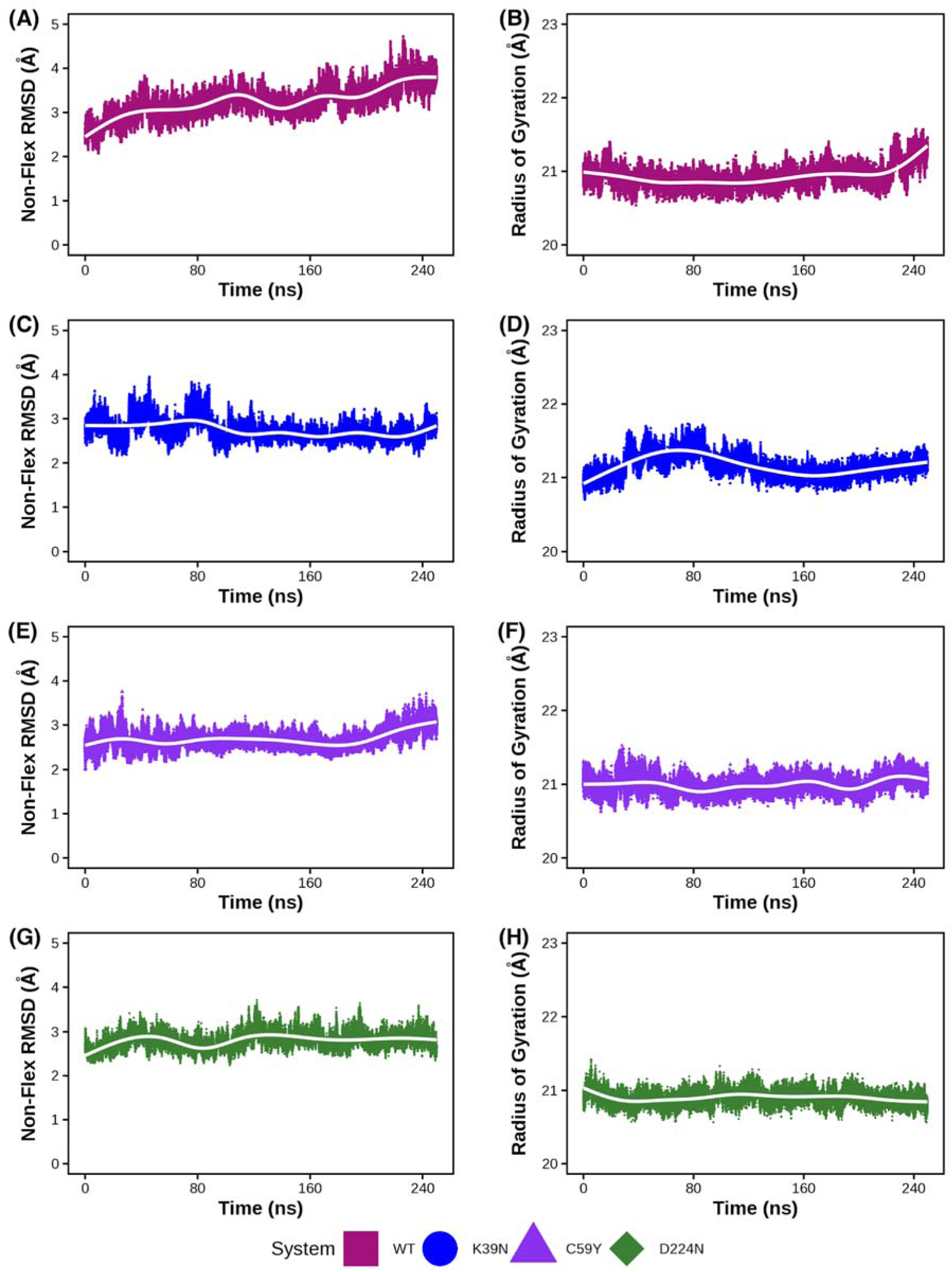
Non-flexible RMSD and radius of gyration. Non-flexible RMSD (left) and radius of gyration (right) of the first MD replicate.

**Supplementary Figure 9.**
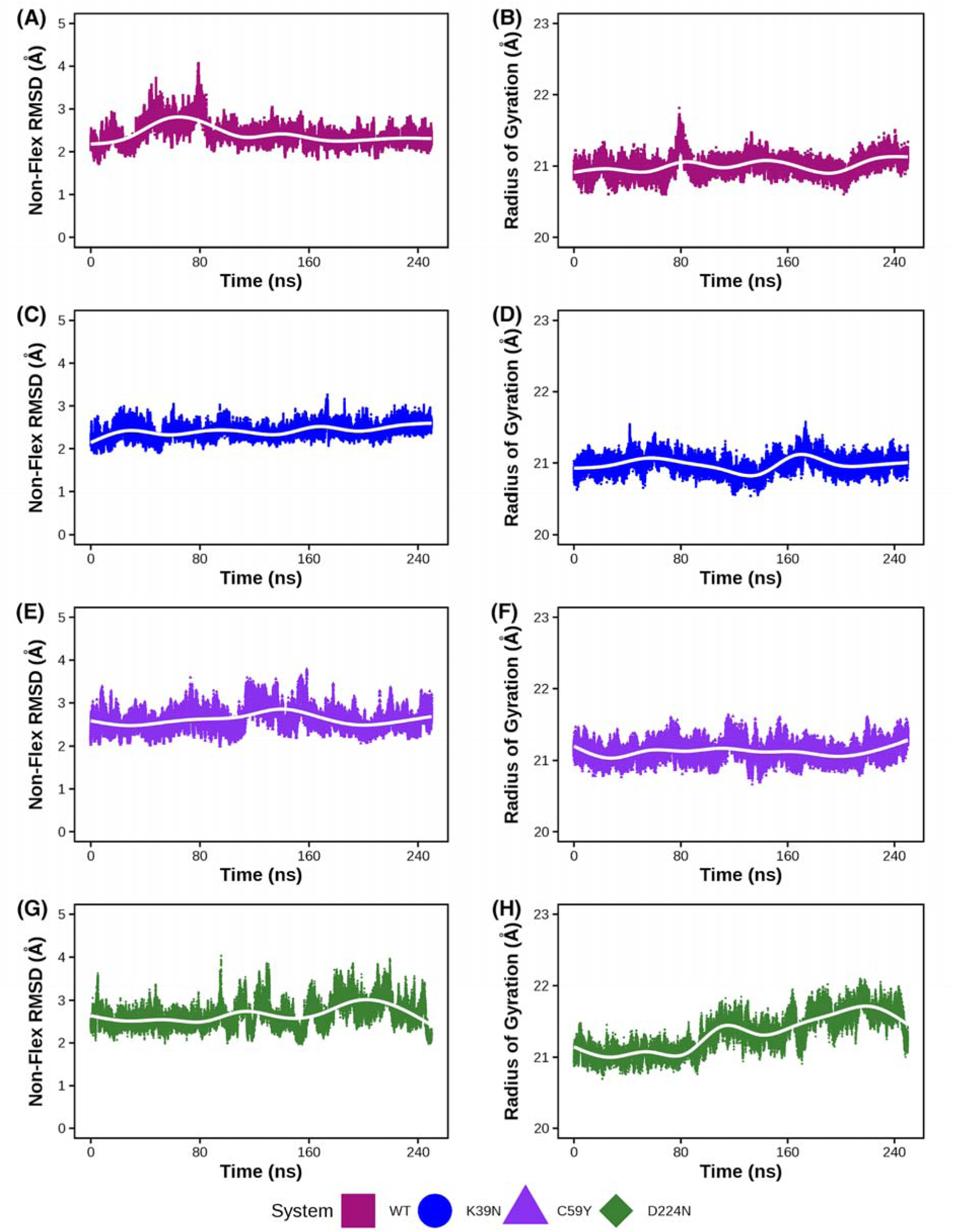
Non-flexible RMSD and radius of gyration. Non-flexible RMSD (left) and radius of gyration (right) of the second MD replicate.

**Supplementary Figure 10.**
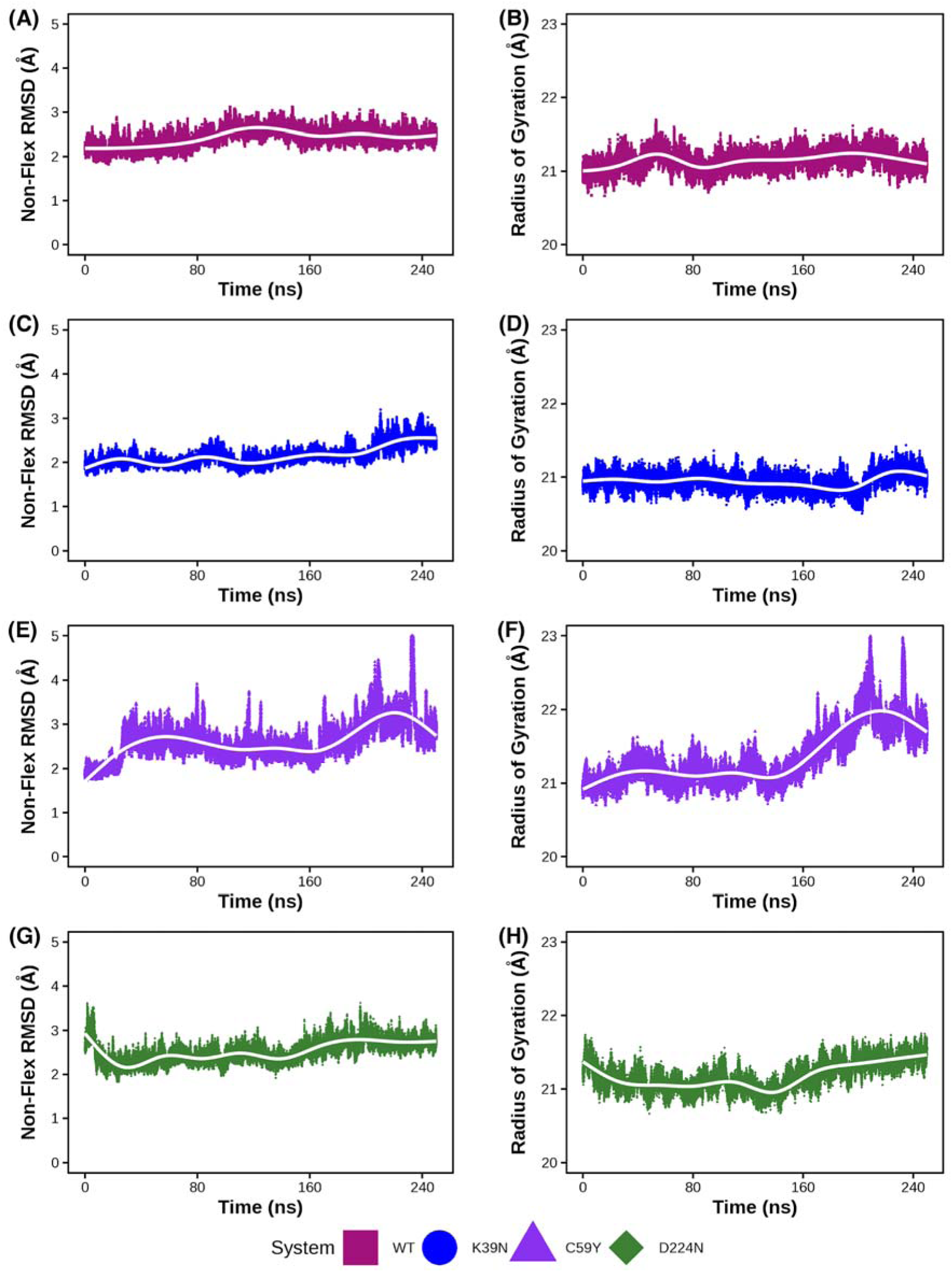
Non-flexible RMSD and radius of gyration. Non-flexible RMSD (left) and radius of gyration (right) of the third MD replicate.

**Supplementary Figure 11.**
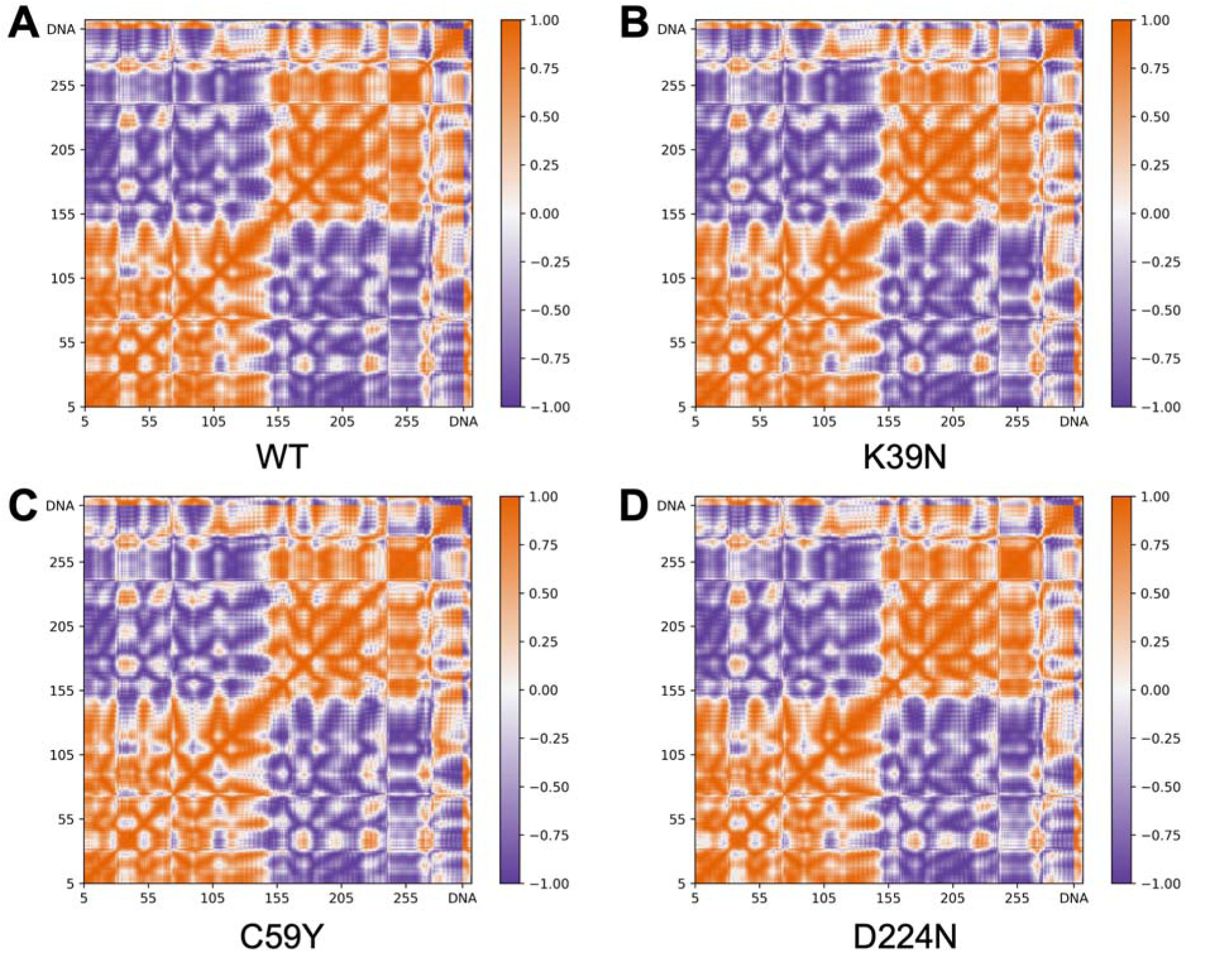
Correlation matrices. Correlation matrices for (A) WT, (B) K39N, (C) C59Y, and (D) D224N. Areas of correlation are orange (1.0), areas with no correlation are white (0.0), and areas with anti-correlation are purple (-1.0).

**Supplementary Figure 12.**
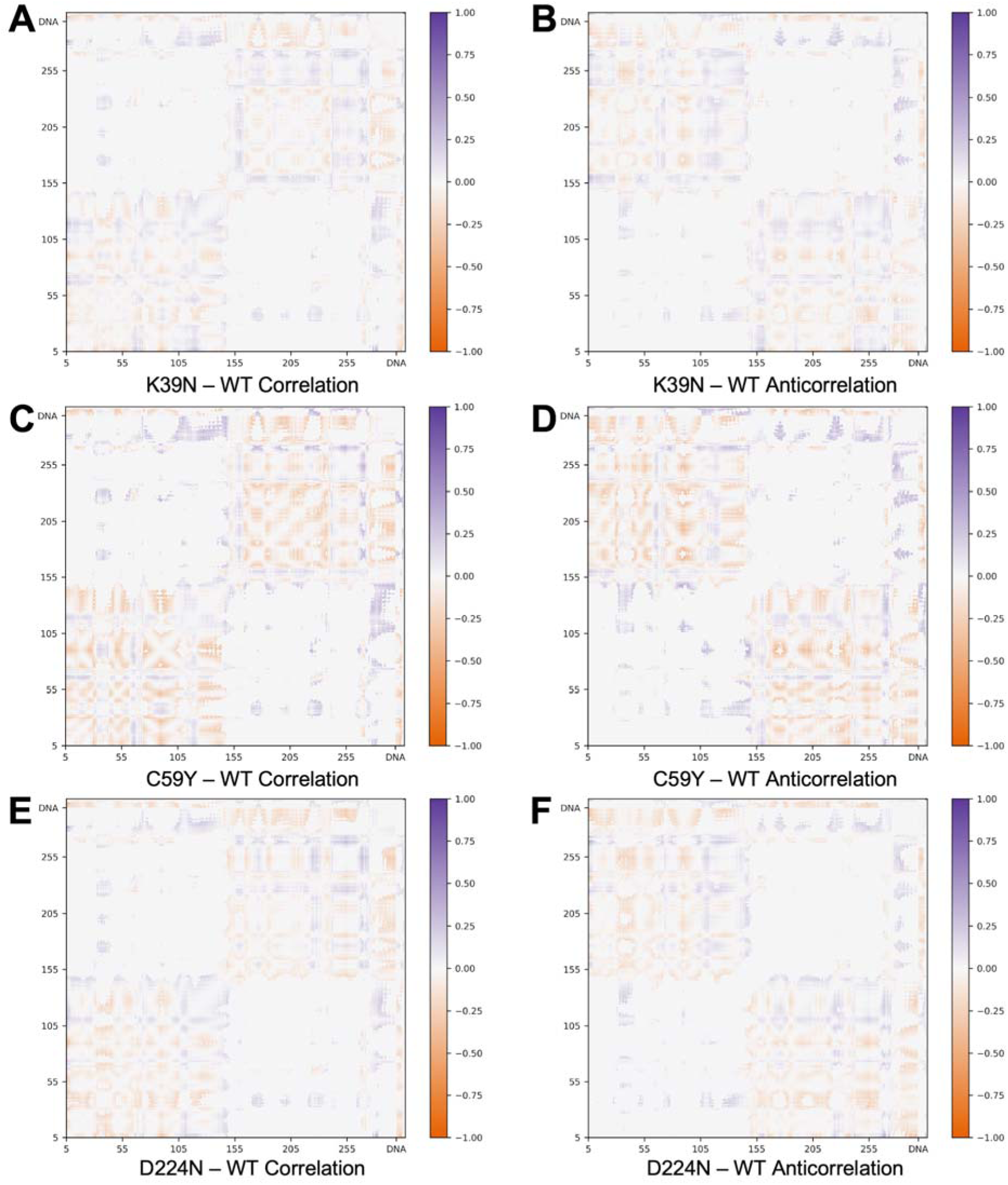
Differences in correlated and anti-correlated motions. Differences in (A,C,E) correlated motions and (B,D,F) anticorrelated motions for (A–B) K39N – WT, (C–D) C59Y – WT, and (E–F) D224N – WT. Areas where that type of motion is more present in WT are orange, and areas where that type of motion is more present in the variant are purple.

**Supplementary Figure 13.**
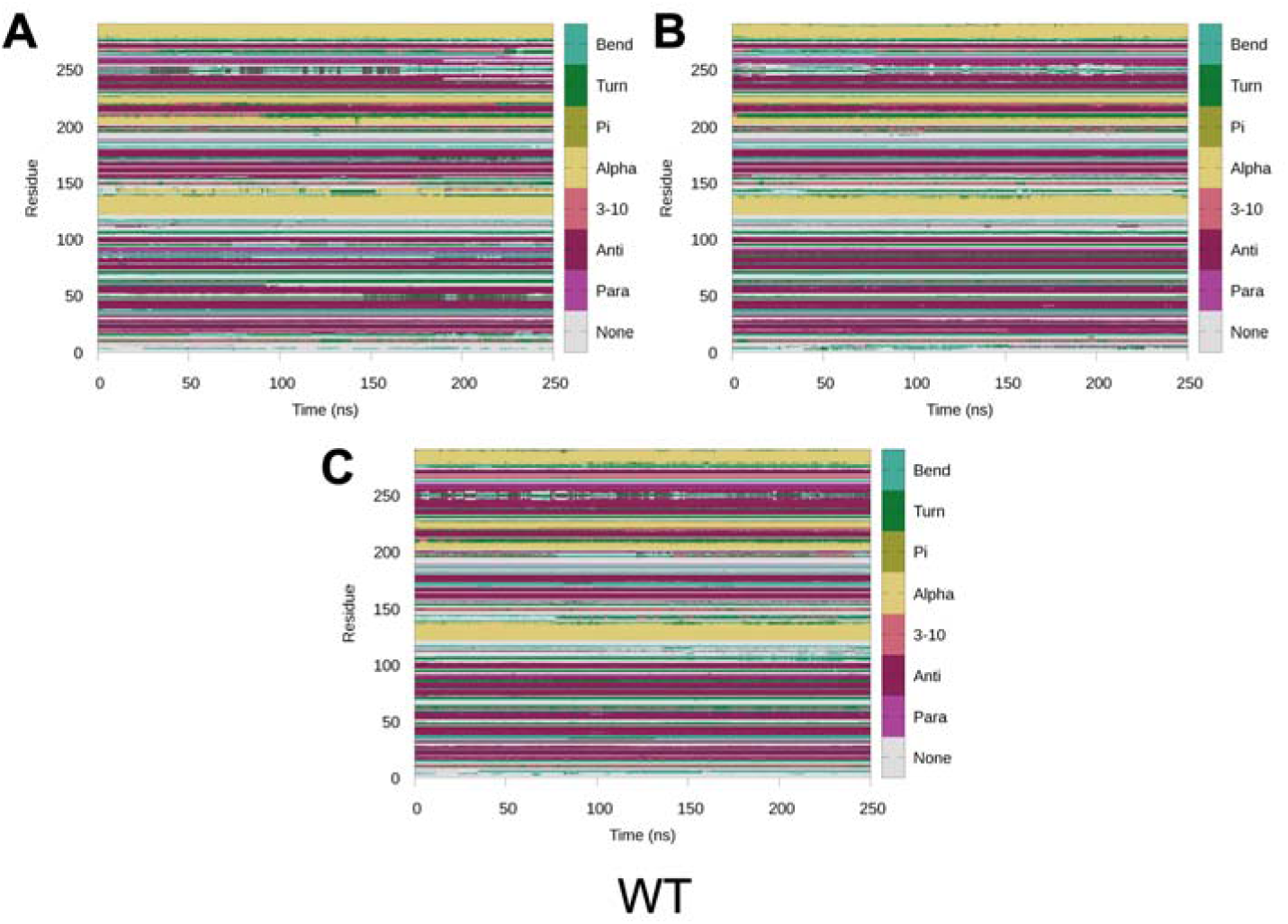
WT secondary structure. Secondary structure analysis of each replicate of WT.

**Supplementary Figure 14.**
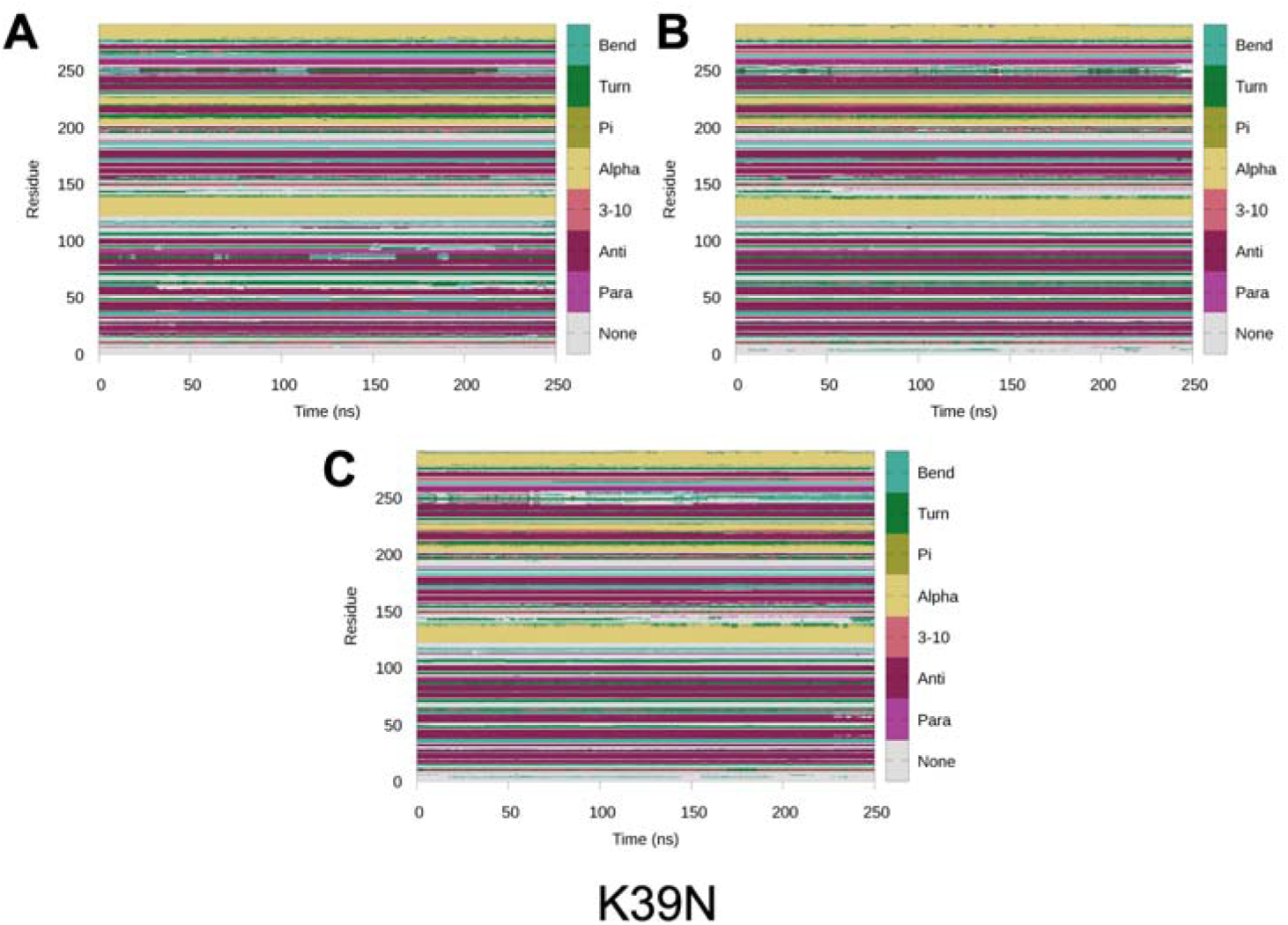
K39N secondary structure. Secondary structure analysis of each replicate of K39N.

**Supplementary Figure 15.**
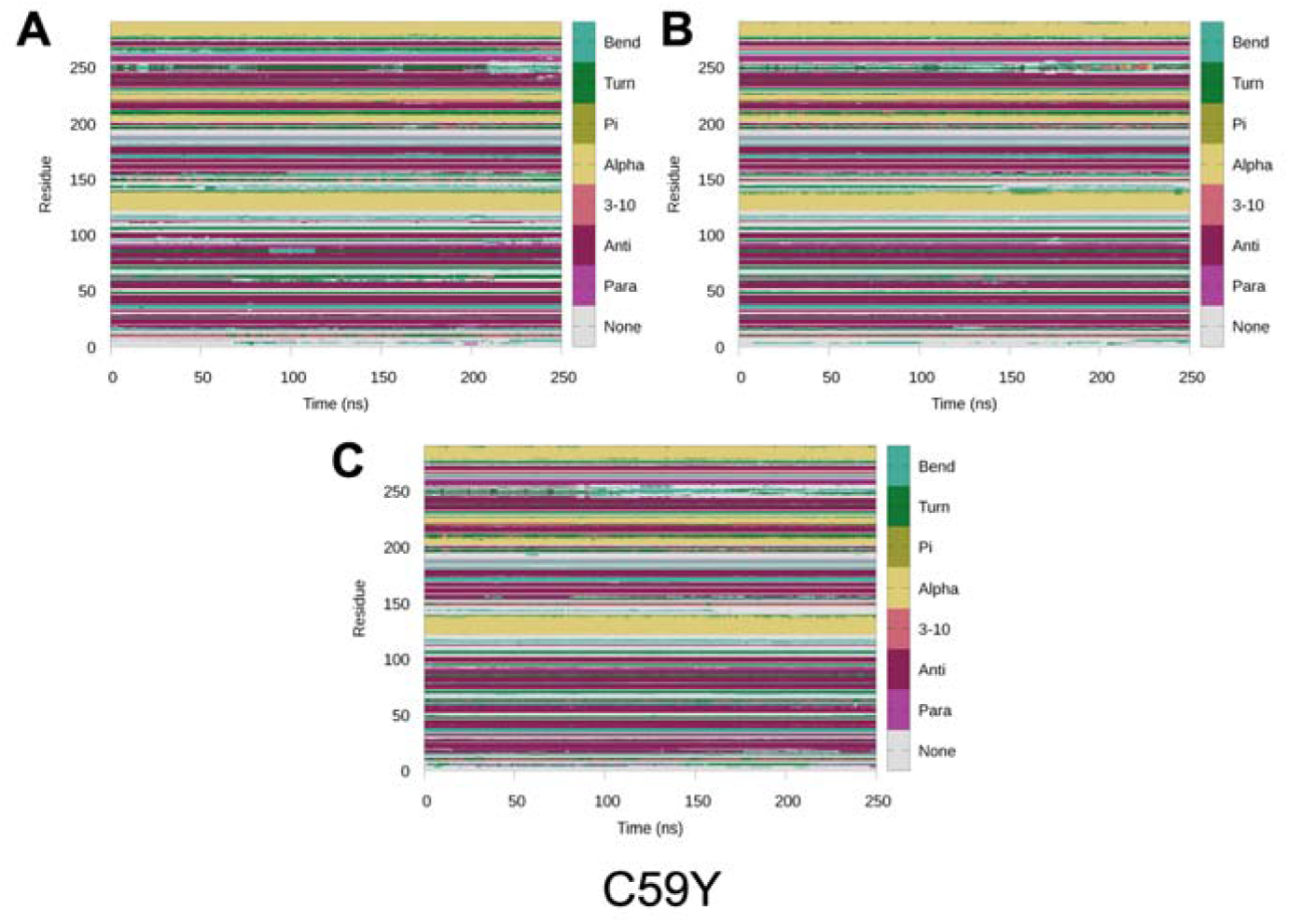
C59Y secondary structure. Secondary structure analysis of each replicate of C59Y.

**Supplementary Figure 16.**
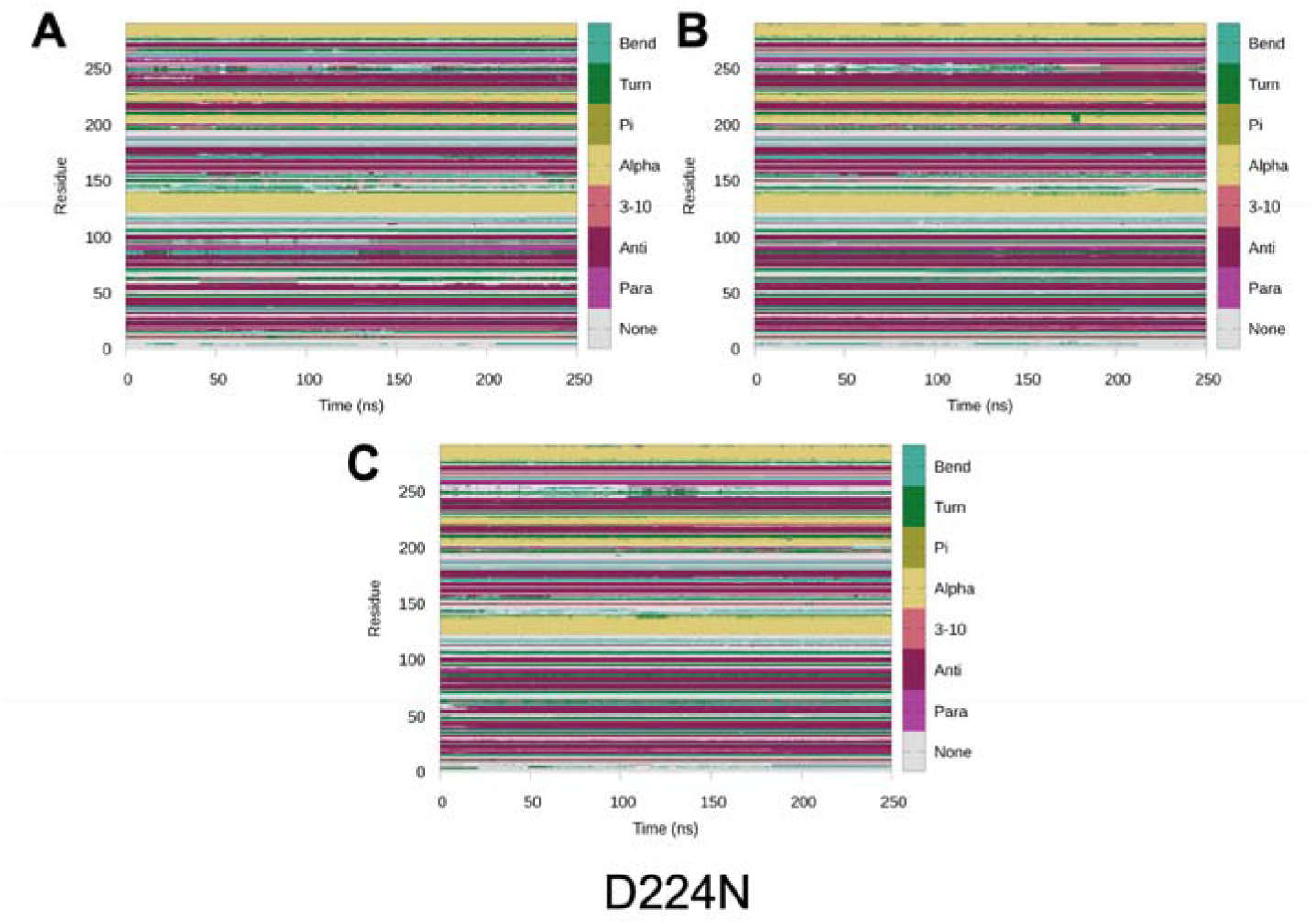
D224N secondary structure. Secondary structure analysis of each replicate of D224N.

**Supplementary Figure 17.**
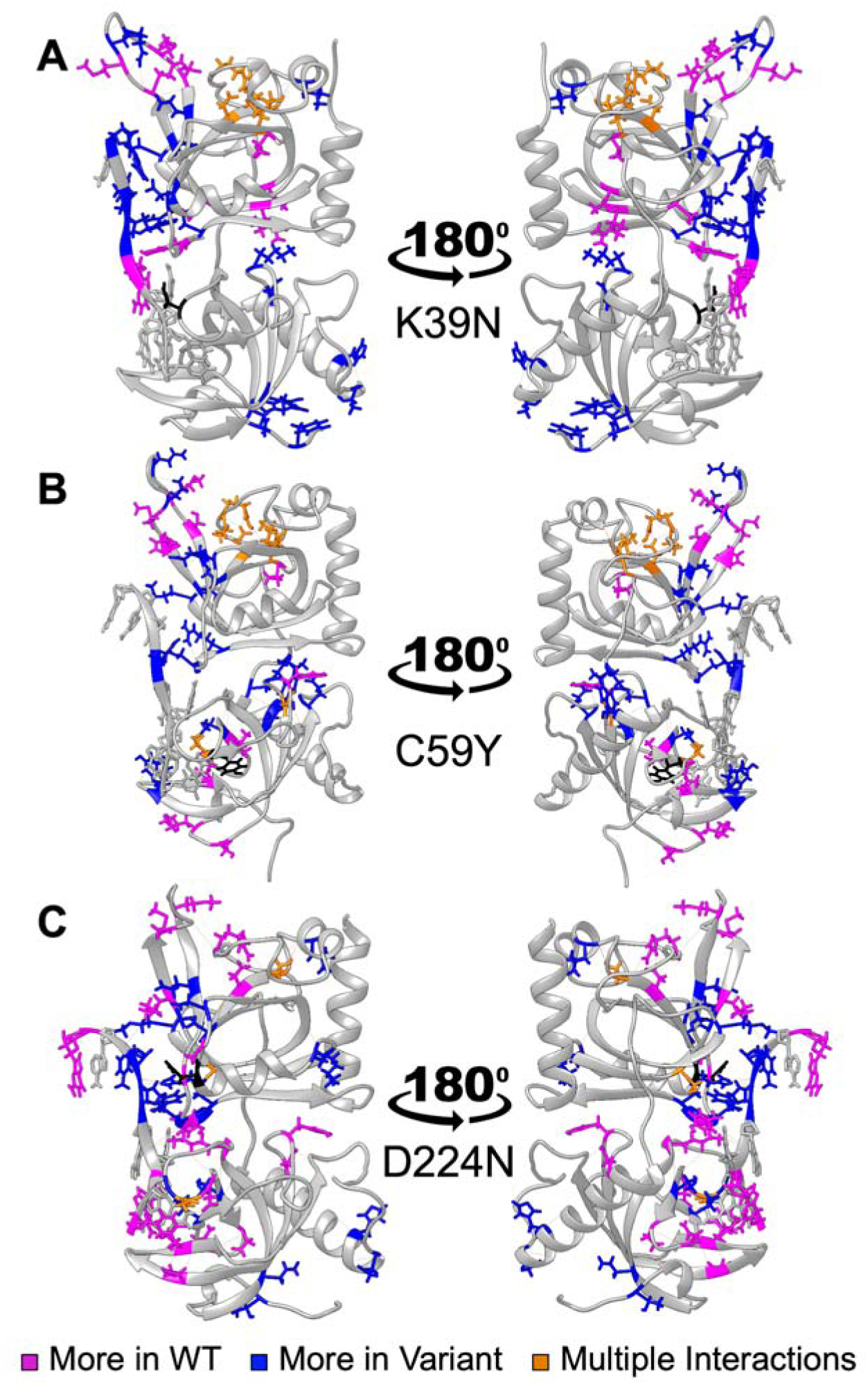
Differences in hydrogen bond interactions. Differences in hydrogen bond interactions greater than 20% of the total simulation time for (A) WT and K39N, (B) WT and C59Y, and (C) WT and D224N. Interactions that are present for a larger percentage of simulation time in the WT are pink, and those more present in the variant structures are blue. Residues that had a combination of hydrogen bond interactions favored between both WT and the variant are shown in orange. The mutation location is shown in black, and all data are shown on the respective variant structure.

**Supplementary Movie.** First and second normal modes of the WT, K39N, C59Y, and D224N POT1-ssDNA variants from molecular dynamics simulations. The mutation position is colored pink. The movie can be found at https://drive.google.com/file/d/1iC_pwzvlFwFdkimvBYM8_8CM_z7mcAXR/view?usp=sharing

## Supplementary Tables and legends

**Supplementary Table 1.**
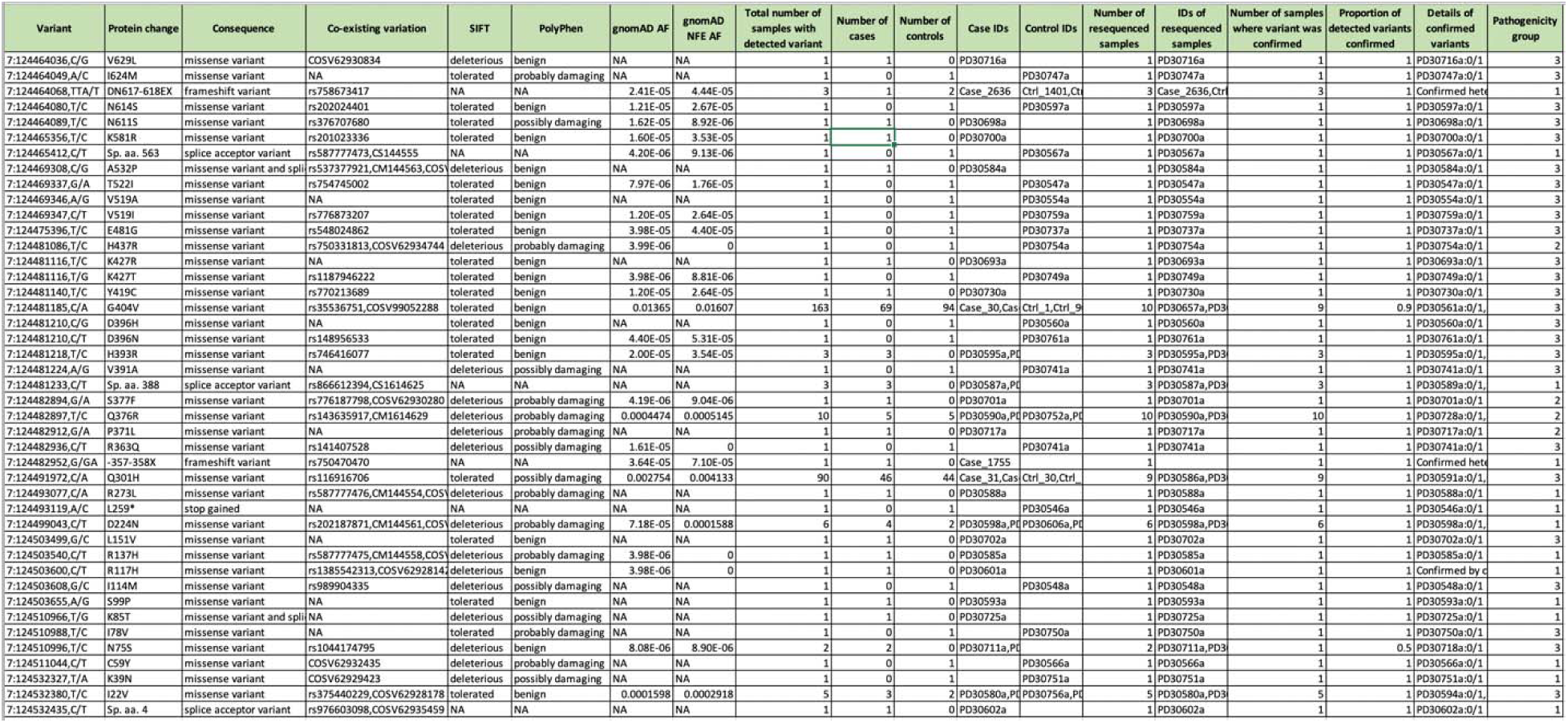
**All protein-altering variants in POT1 found in this study.** Coordinates are in the GRCh37 reference genome. Consequences were predicted with VEP, Ensembl release 104, which also annotated co-existing variation, SIFT and PolyPhen pathogenicity predictions, and allele frequencies in the gnomAD database. All variants in this table were confirmed by resequencing of the original samples by a different method.

**Supplementary Table 2.**
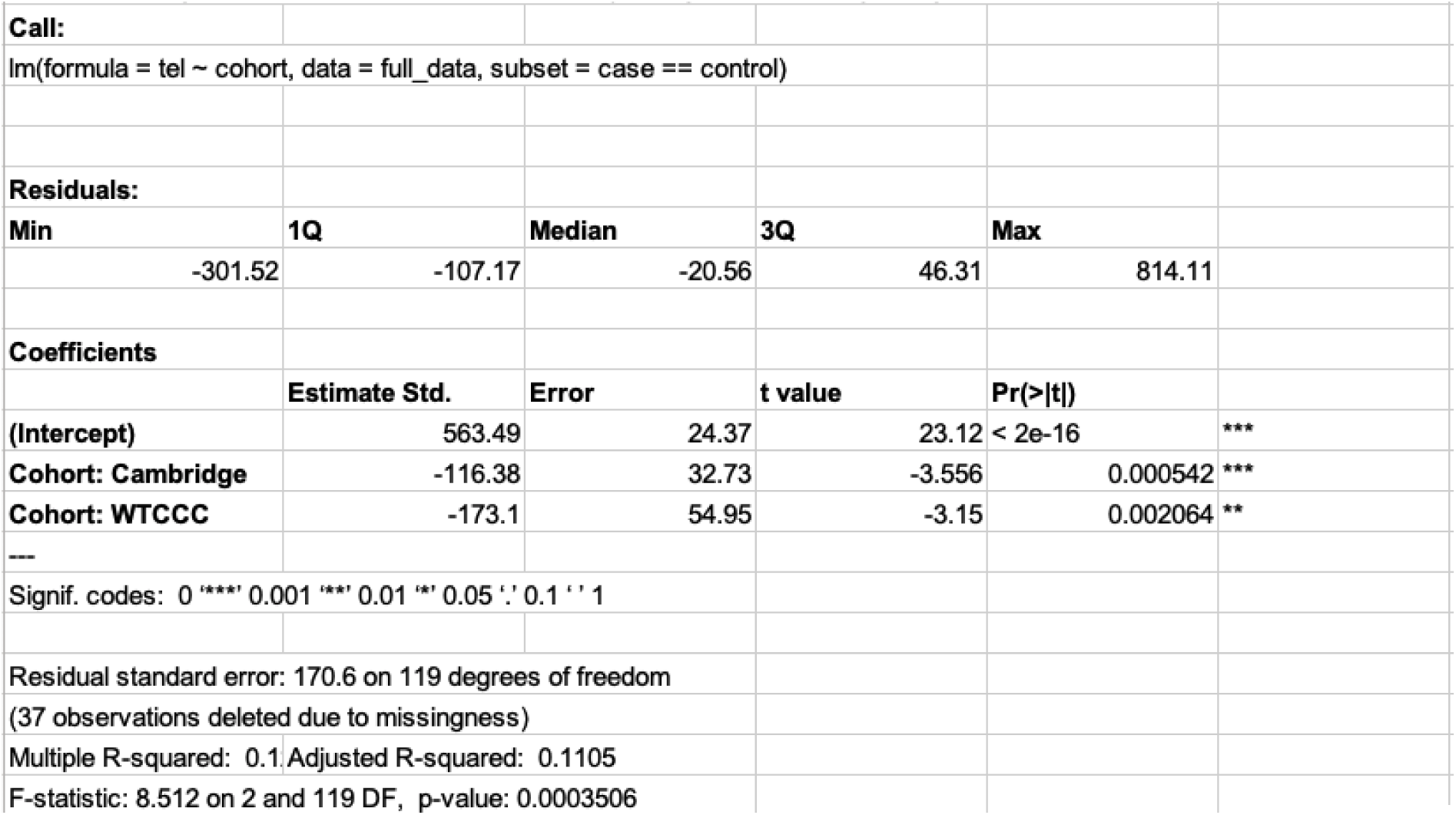
**Linear model used for adjusting telomere lengths by cohort.**

**Supplementary Table 3.** **Change in interactions with K39N.** Residues that showed the greatest differences (K39N – WT) across systems in their total interaction energy with respect to the K39N residue. The average total interaction energy ± average standard deviation is provided. All values are in kcal mol^-1^.

|  | K39N – WT |
| --- | --- |
| Ala6 | -7.42 ± 0.01 |
| Lys18 | -9.83 ± 0.02 |
| Lys30 | -11.88 ± 0.02 |
| Lys33 | -25.84 ± 0.11 |
| Pro34 | 1.05 ± 0.01 |
| Leu37 | 2.27 ± 0.04 |
| Asp42 | 25.20 ± 0.02 |
| Asp51 | 9.96 ± 0.01 |
| Lys56 | -13.17 ± 0.01 |
| Glu67 | 13.36 ± 0.09 |
| Lys74 | -10.33 ± 0.05 |
| Asp77 | 11.35 ± 0.04 |
| Arg80 | -9.92 ± 0.01 |
| Arg83 | -10.83 ± 0.03 |
| Lys85 | -14.64 ± 0.04 |
| Lys90 | -20.02 ± 0.04 |
| Lys91 | -13.28 ± 0.08 |
| Glu92 | 13.97 ± 0.08 |
| Glu107 | 9.75 ± 0.04 |
| Arg117 | -10.99 ± 0.05 |
| Lys121 | -9.29 ± 0.04 |
| Glu128 | 7.61 ± 0.01 |
| Asp129 | 9.00 ± 0.01 |
| Lys131 | -7.24 ± 0.01 |
| Glu134 | 7.64 ± 0.01 |
| Arg137 | -8.59 ± 0.03 |
| Lys152 | -8.87 ± 0.05 |
| Asp155 | 8.37 ± 0.03 |
| Asp163 | 12.76 ± 0.05 |
| Lys171 | -13.01 ± 0.17 |
| Glu173 | 14.63 ± 0.24 |
| Asp175 | 15.10 ± 0.25 |
| Lys182 | -9.87 ± 0.08 |
| Asp185 | 9.23 ± 0.05 |
| Arg188 | -7.60 ± 0.04 |
| Arg195 | -9.10 ± 0.04 |
| Asp200 | 8.91 ± 0.08 |
| Glu204 | 10.90 ± 0.12 |
| Asp206 | 8.41 ± 0.06 |
| Arg212 | -7.42 ± 0.05 |
| Asp219 | 10.27 ± 0.08 |
| Asp224 | 25.66 ± 0.78 |
| Arg231 | -17.86 ± 0.34 |
| Lys234 | -10.94 ± 0.12 |
| Arg240 | -11.45 ± 0.08 |
| Lys247 | -11.71 ± 0.06 |
| Glu254 | 7.68 ± 0.09 |
| Glu262 | 11.20 ± 0.07 |
| Gly268 | -1.31 ± 0.04 |
| Arg273 | -18.62 ± 0.17 |
| Arg276 | -14.51 ± 0.08 |
| Glu280 | 9.27 ± 0.06 |
| Asp284 | 9.78 ± 0.04 |
| Asp286 | 8.08 ± 0.04 |
| Lys289 | $-8.51 \pm 0.05$ |
| Lys290 | $-7.54 \pm 0.03$ |
| Asp291 | $8.09 \pm 0.04$ |
| Glu293 | $7.17 \pm 0.03$ |
| Ala299 | $6.71 \pm 0.03$ |
| dT1 | $9.38 \pm 0.14$ |
| dT2 | $32.79 \pm 0.71$ |
| dA3 | $47.18 \pm 1.32$ |
| dG4 | $27.20 \pm 0.46$ |
| dG5 | $17.45 \pm 0.28$ |
| dG6 | $18.03 \pm 0.38$ |
| dT7 | $15.12 \pm 0.29$ |
| dT8 | $17.25 \pm 0.41$ |
| dA9 | $19.70 \pm 0.82$ |
| dG10 | $12.51 \pm 0.59$ |

**Supplementary Table 4.** **Change in interactions with D224N.** Residues that showed the greatest differences (D224N – WT) across systems in their total interaction energy with respect to the D224N residue. The average total interaction energy ± average standard deviation is provided. All values are in kcal mol^-1^.

|  | D224N – WT |
| --- | --- |
| Ala6 | 6.59 ± 0.02 |
| Lys18 | 7.97 ± 0.05 |
| Lys30 | 11.83 ± 0.03 |
| Lys33 | 23.01 ± 0.11 |
| Leu37 | -1.06 ± 0.05 |
| Lys39 | 26.00 ± 0.79 |
| Asp42 | -15.03 ± 0.19 |
| Asp51 | -9.06 ± 0.02 |
| Lys56 | 12.19 ± 0.03 |
| Glu67 | -12.81 ± 0.11 |
| Lys74 | 11.63 ± 0.02 |
| Asp77 | -11.84 ± 0.01 |
| Arg80 | 8.79 ± 0.02 |
| Arg83 | 8.53 ± 0.04 |
| Lys85 | 9.92 ± 0.06 |
| Lys90 | 14.16 ± 0.11 |
| Lys91 | 10.66 ± 0.06 |
| Glu92 | -11.49 ± 0.04 |
| Glu107 | -10.35 ± 0.01 |
| Arg117 | 11.35 ± 0.03 |
| Lys121 | 8.55 ± 0.05 |
| Glu128 | -7.02 ± 0.01 |
| Asp129 | -8.20 ± 0.02 |
| Lys131 | 6.78 ± 0.01 |
| Glu134 | -7.63 ± 0.01 |
| Arg137 | 8.72 ± 0.01 |
| Lys152 | 11.27 ± 0.02 |
| Asp155 | -10.60 ± 0.03 |
| Asp163 | -16.07 ± 0.04 |
| Lys171 | 18.41 ± 0.02 |
| Glu173 | -24.24 ± 0.09 |
| Val174 | 1.31 ± 0.01 |
| Asp175 | -32.49 ± 0.33 |
| Ala177 | -4.17 ± 0.21 |
| Ser178 | 1.19 ± 0.05 |
| Lys182 | 15.15 ± 0.06 |
| Asp185 | -12.22 ± 0.02 |
| Arg188 | 10.21 ± 0.03 |
| Arg195 | 13.68 ± 0.08 |
| Asp200 | -12.76 ± 0.06 |
| Glu204 | -14.84 ± 0.01 |
| Asp206 | -11.05 ± 0.01 |
| Arg212 | 9.52 ± 0.02 |
| Asp219 | -16.37 ± 0.08 |
| His226 | 3.76 ± 0.09 |
| Val227 | 3.45 ± 0.07 |
| His228 | 3.29 ± 0.14 |
| Val229 | 1.34 ± 0.01 |
| Ala230 | 1.42 ± 0.00 |
| Arg231 | $29.38 \pm 0.11$ |
| Lys234 | $12.94 \pm 0.03$ |
| Arg240 | $13.76 \pm 0.03$ |
| Lys247 | $19.67 \pm 0.21$ |
| Glu254 | $-10.82 \pm 0.09$ |
| Glu262 | $-19.03 \pm 0.12$ |
| Leu265 | $-1.88 \pm 0.03$ |
| His266 | $1.46 \pm 0.21$ |
| Gly267 | $5.74 \pm 0.12$ |
| Arg273 | $30.99 \pm 0.18$ |
| Arg276 | $15.50 \pm 0.07$ |
| Glu280 | $-10.55 \pm 0.01$ |
| Asp284 | $-11.26 \pm 0.01$ |
| Asp286 | $-9.21 \pm 0.00$ |
| Lys289 | $9.88 \pm 0.00$ |
| Lys290 | $8.73 \pm 0.01$ |
| Asp291 | $-9.95 \pm 0.02$ |
| Glu293 | $-8.66 \pm 0.01$ |
| Ala299 | $-8.51 \pm 0.02$ |
| dT1 | $-4.29 \pm 0.11$ |
| dT2 | $-14.74 \pm 0.37$ |
| dA3 | $-18.74 \pm 0.52$ |
| dG4 | $-17.86 \pm 0.36$ |
| dG5 | $-14.14 \pm 0.21$ |
| dG6 | $-17.65 \pm 0.27$ |
| dT7 | $-17.94 \pm 0.21$ |
| dT8 | $-24.81 \pm 0.39$ |
| dA9 | $-28.25 \pm 0.53$ |
| dG10 | $-10.10 \pm 1.25$ |

**Supplementary Table 5.** **Change in interactions with dG6 for K39N.** Residues that showed the greatest differences (K39N – WT) across systems in their total interaction energy with respect to the dG6 residue. The average total interaction energy ± average standard deviation is provided. All values are in kcal mol^-1^.

|  | K39N – WT |
| --- | --- |
| Lys30 | 2.41 ± 0.76 |
| Lys33 | 11.95 ± 3.73 |
| Lys39 | 18.03 ± 0.38 |
| Lys56 | 4.10 ± 1.02 |
| Lys74 | 1.11 ± 0.35 |
| Asp77 | -1.26 ± 0.37 |
| Lys91 | 1.13 ± 0.69 |
| Glu92 | 4.95 ± 0.75 |
| Asp163 | -2.28 ± 0.56 |
| Arg240 | 2.04 ± 0.43 |
| Thr269 | -1.25 ± 0.23 |
| Ser270 | 1.06 ± 0.75 |
| Tyr271 | 1.59 ± 0.51 |
| Gly272 | 1.14 ± 0.18 |
| Arg273 | 2.47 ± 1.10 |
| Asp284 | -1.07 ± 0.25 |
| dA9 | -2.43 ± 0.39 |

**Supplementary Table 6.** **Change in interactions with dG6 for C59Y.** Residues that showed the greatest differences (C59Y – WT) across systems in their total interaction energy with respect to the dG6 residue. The average total interaction energy ± average standard deviation is provided. All values are in kcal mol^-1^.

|  | C59Y – WT |
| --- | --- |
| Lys18 | -1.01 ± 0.19 |
| Lys33 | 8.00 ± 2.67 |
| Asp51 | 1.28 ± 0.20 |
| Lys56 | -10.02 ± 0.80 |
| Lys90 | -2.62 ± 0.66 |
| Glu92 | 5.88 ± 0.76 |
| Asp163 | 1.31 ± 0.36 |
| Asp224 | 1.13 ± 0.41 |
| Lys247 | -1.43 ± 0.23 |
| Thr269 | -1.09 ± 0.27 |
| Ser270 | -1.09 ± 0.50 |
| Arg273 | -1.88 ± 0.83 |
| Arg276 | -1.37 ± 0.25 |
| dA3 | -1.31 ± 0.29 |
| dG4 | -3.12 ± 0.70 |
| dT8 | 1.02 ± 0.38 |
| dA9 | -1.94 ± 0.40 |

**Supplementary Table 7.** **Change in interactions with dG6 for D224N.** Residues that showed the greatest differences (D224N – WT) across systems in their total interaction energy with respect to the dG6 residue. The average total interaction energy ± average standard deviation is provided. All values are in kcal mol^-1^.

|  | D224N – WT |
| --- | --- |
| Lys30 | 1.71 ± 0.94 |
| Lys33 | 11.17 ± 3.92 |
| Lys39 | 2.07 ± 3.92 |
| Asp42 | -1.02 ± 0.49 |
| Lys56 | 3.74 ± 1.13 |
| Glu67 | -1.26 ± 0.33 |
| Lys74 | 1.13 ± 0.41 |
| Asp77 | -1.21 ± 0.45 |
| Lys90 | 1.76 ± 0.63 |
| Lys91 | 2.06 ± 0.85 |
| Glu92 | 4.17 ± 1.03 |
| Asp163 | -1.70 ± 0.77 |
| Asp224 | -17.65 ± 0.27 |
| Arg240 | 1.72 ± 0.54 |
| Tyr271 | 1.78 ± 0.49 |
| Gly272 | 1.26 ± 0.18 |
| Arg273 | 2.96 ± 1.27 |
| dA3 | -1.56 ± 0.43 |
| dG4 | -5.10 ± 0.99 |
| dT8 | 1.11 ± 0.33 |

**Supplementary Table 8. Samples included in this study.** The list shows all 6,227 samples in this study, with their ID, proportion of high-quality bases sequenced in the 1st sequencing round, case/control status and cohort of origin. *This is a long table and can be found at https://www.dropbox.com/s/2nb3f7sht0pevjr/Supplementary_Table_3.xlsx?dl=0

**Supplementary Table 9. All protein-altering variants in POT1 found in this study after the first round of sequencing.** Coordinates are in the GRCh37 reference genome. Details of the number of samples that were re-sequenced for confirmation and other information are included. Variants confirmed are highlighted in green. *This is a long table and can be found at https://www.dropbox.com/s/z4830tuwp1umaj4/Supplementary_Table_4.xlsx?dl=0

**Supplementary Table 10. List of samples that were re-sequenced by Illumina fo confirmation.** The average coverage of coding *POT1* exons is included. In red, samples with a coverage lower than 10. *This is a long table and can be found at https://www.dropbox.com/s/wn13y70iyfsnmi9/Supplementary_Table_5.xlsx?dl=0

**Supplementary Table 11.** **List of samples that were re-sequenced by capillary.** Information about the variants re-sequenced and the result of the experiment is included. In green, samples and variants that were confirmed.

| Sample | Variant tested | Result |
| --- | --- | --- |
| PD30582a | C307R | Not present |
| PD30601a | R117H | Present |
| PD30604a | H393R | Present |
| PD30608a | I22V | Present |
| PD30610a | P601L | Not present |
| PD30624a | E92K, R83G | Not present |
| PD30681a | V391I | Not present |
| PD30687a | Q376* | Not present |
| PD30692a | W194* | Not present |
| PD30696a | P34S | Not present |
| Case_2636 | DN617-618EX | Present |
| Ctrl_1401 | DN617-618EX | Present |
| Ctrl_3151 | DN617-618EX | Present |
| Case_1755 | -357-358X | Present |
| Ctrl_2641 | E204* | Not present |
| Ctrl_2500 | D200N | Not present |
| Case_2329 | D185E | Not present |
| Ctrl_2165 | D185E | Not present |
| Case_1223 | D175* | Not present |

**Supplementary Table 12.** **List of samples with variants in known melanoma predisposition genes.** Genes *CDK4*, *CDKN2A* and *BAP1* were checked for variants. Only samples that were resequenced by Illumina (Supplementary Table 3) were assessed.

| Sample | *POT1* variant | Variant in pathogenic gene |
| --- | --- | --- |
| PD30744a | Q376R | *CDKN2A* A148T |
| PD30549a | G404V | *CDKN2A* A148T |
| PD30730a | G404V & Y419C | *CDKN2A* A60T |

